# Burden of influenza hospitalization among high-risk groups in the United States

**DOI:** 10.1101/2021.12.10.21267528

**Authors:** Aimee Near, Jenny Tse, Yinong Young-Xu, David K. Hong, Carolina M. Reyes

## Abstract

**Background:** Seasonal influenza poses a substantial clinical and economic burden in the United States and vulnerable populations, including the elderly and those with comorbidities, are at elevated risk for influenza-related medical complications.

**Methods:** We conducted a retrospective cohort study using the IQVIA PharMetrics® Plus claims database in two stages. In Stage 1, we identified patients with evidence of medically-attended influenza during influenza seasons from October 1, 2014 to May 31, 2018 (latest available data for Stage 1) and used a multivariable logistic regression model to identify patient characteristics that predicted 30-day influenza-related hospitalization. Findings from Stage 1 informed high-risk subgroups of interest for Stage 2, where we selected cohorts of influenza patients during influenza seasons from October 1, 2014 to March 1, 2019 and used 1:1 propensity score matching to patient without influenza with similar high-risk characteristics to compare influenza-attributable rates of all-cause hospital and emergency department visits during follow-up (30-day and in index influenza season).

**Results:** In Stage 1, more than 1.6 million influenza cases were identified, of which 18,509 (1.2%) had a hospitalization. Elderly age was associated with 9 times the odds of hospitalization (≥65 years vs. 5-17 years; OR=9.4, 95% CI 8.8-10.1) and select comorbidities were associated with 2-3 times the odds of hospitalization. In Stage 2, elderly influenza patients with comorbidities had 3 to 7 times higher 30-day hospitalization rates compared to matched patients without influenza, including patients with congestive heart failure (41.0% vs.7.9%), chronic obstructive pulmonary disease (34.6% vs. 6.1%), coronary artery disease (22.8% vs. 3.8%), and late-stage chronic kidney disease (44.1% vs. 13.1%; all p<0.05).

**Conclusions:** The risk of influenza-related complications is elevated in the elderly, especially those with certain underlying comorbidities, leading to excess healthcare resource utilization. Continued efforts, beyond currently available vaccines, are needed to reduce influenza burden in high-risk populations.

## 1. Introduction

Despite the availability of vaccines, the burden of seasonal influenza remains high in the United States (US), contributing to excess morbidity, mortality, and healthcare resource utilization (HRU). The Centers for Disease Control and Prevention (CDC) estimates that influenza accounted for 4.3-21 million medical visits, 140,000-810,000 hospitalizations, and 12,000-61,000 deaths annually in the US during the 2010-11 through 2019-20 influenza seasons [1]. In turn, the estimated total economic burden of influenza is substantial at $11.2 billion (ranging from $6.3-$25.3 billion) [2] and as high as $87.1 billion (95% confidence interval [CI], $47.2-$149.5) [3]. Direct medical costs have been estimated at $3.2 billion annually, of which 70% ($2.3 billion) is due to hospitalizations [2], despite hospitalization in only 1-2% of medically-attended influenza cases [1].

Although influenza is generally self-limiting with mild symptoms in healthy individuals [4], certain vulnerable populations are at elevated risk for serious influenza-related medical complications. For example, while the elderly population ≥65 years of age has the lowest median incidence of influenza (3.9%) compared to children 0-17 years (9.3%) or adults 18-64 years (8.8%) [5], they account for 50-70% of influenza-related hospitalizations, 70-85% of deaths [6], and 42.7% of direct medical costs [2]. Chronic medical conditions, including pulmonary, cardiovascular, renal, hepatic, and metabolic disorders, have also been identified as predictors of influenza-related complications [7-12]. Vaccination is recognized as the most effective prevention strategy for seasonal influenza, but in the 2019-2020 season, only 51.8% of persons six months or older were vaccinated [13].

As both vaccination against influenza infections and treatment for complications evolve, there is limited up-to-date research quantifying risk factors associated with hospitalization or the added economic burden attributable to seasonal influenza among patients vulnerable to complications using real-world data. To that end, this study aimed to identify risk factors for influenza-related hospitalization (Stage 1) and to evaluate the economic burden of influenza in at-risk elderly populations (Stage 2).

## 2. Methods

### 2.1. Study data source

The data source for this retrospective cohort study was IQVIA PharMetrics^®^ Plus, a health plan claims database comprised of fully adjudicated medical and pharmacy claims from more than 100 commercial health plans, covering more than 150 million unique enrollees representative of the commercially insured US population. All data are Health Insurance Portability and Accountability Act (HIPAA)-compliant to protect patient privacy.[14, 15] As this retrospective cohort analysis was conducted using de-identified HIPAA-compliant data, Institutional Review Board (IRB) review was not required for this study.

### 2.2. Study period and population

This study was conducted in two stages, with separate patient selection criteria in each stage. Briefly, in Stage 1, patients with evidence of ≥1 influenza diagnosis (i.e. “medically-attended influenza patients”) were identified in the database from October 1, 2014 to May 31, 2018, a timeframe that includes the four most recent influenza seasons at the time of data extraction. The index date was the date of the earliest observed influenza diagnosis occurring during an influenza season (spanning from October 1 to May 31 of the subsequent year). Patients were required to have ≥12 months of continuous enrollment in their health plan before index (baseline) and ≥30 days after index (follow-up) and either index influenza diagnosis in the primary position or evidence of influenza lab test order ±14 days of index. Influenza diagnoses were identified using International Classification of Diseases, 9^th^ and 10^th^ revision (ICD-9/10) codes (ICD-9: 487-488; ICD-10: J09-J11). Influenza patients who met all inclusion and exclusion criteria were stratified into two mutually exclusive groups (hospitalized vs. non-hospitalized) based on the presence of ≥1 influenza-related hospitalization. Influenza-related hospitalization was defined as an inpatient visit with a diagnosis code for influenza or an influenza-related complication (respiratory, renal, cardiovascular, neurological/musculoskeletal, and other conditions [e.g., conjunctivitis, dehydration, liver inflammation/hepatitis, sepsis]) associated with the claim in any position within 30 days after the index influenza diagnosis [16-21].

In Stage 2, patients who were ≥65 years of age were identified during the study period from October 1, 2013 to March 31, 2019 and then selection criteria were applied separately to the influenza cohort and the non-influenza cohort in order to compare influenza-attributable burden. Selection criteria for the influenza cohort were similar to those applied in Stage 1 and the index date was the date of the earliest influenza diagnosis occurring during an influenza season from October 1, 2014 to March 1, 2019. Patients in the non-influenza comparator cohort had no evidence of influenza diagnosis during the study period and their index dates were assigned to replicate the distribution of index dates observed in the influenza cohort. The elderly influenza and non-influenza cohorts were further stratified into 12 non-mutually exclusive high-risk subgroups (broadly categorized as pulmonary disease, cardiovascular disease, or renal disease) characterized by baseline comorbidities. Patients were required to have evidence of ≥1 inpatient or ≥2 outpatient diagnoses during the 12-month baseline period for any of the following high-risk subgroups: asthma, chronic obstructive pulmonary disease (COPD), chronic pulmonary disease, atherosclerosis, coronary artery disease (CAD), congestive heart failure (CHF), stroke, valvular disease, old myocardial infarction (MI; defined using diagnosis codes indicating history of MI), acute MI, late stage chronic kidney disease (CKD; including CKD stage 5, end-stage renal disease, or dialysis), and early stage CKD (including CKD stages 3 or 4). These subgroups were selected based on results from Stage 1 analyses [22], which investigated 29 comorbidities from the Agency for Healthcare Research and Quality (AHRQ) comorbidity software [23] and 10 chronic medical conditions identified by the CDC as increasing the risk for serious influenza-related complications [24].

Within each of the 12 high-risk subgroups, the influenza and non-influenza cohorts were matched using propensity score (PS) matching at a 1:1 ratio with a greedy nearest-neighbor matching algorithm, without replacement and using caliper widths of 0.1 of the standard deviation of the logit of the PS. The logistic regression model to generate the PS included patient demographics, baseline clinical characteristics, and baseline all-cause healthcare costs (USD; detailed list of variables available in **Supplementary Table 1**).

### 2.3. Study measures and statistical analysis

#### Stage 1

Patient demographic and baseline clinical characteristics were assessed during the 12-month baseline period. Comorbidities of interest were derived from 29 comorbidities from the AHRQ comorbidity software [23] and 10 chronic medical conditions identified by the CDC as increasing the risk for serious influenza-related complications [24]. Influenza vaccination was defined as ≥1 National Drug Code (NDC) or Healthcare Common Procedure Coding System (HCPCS) code for influenza vaccination occurring during the index influenza season and prior to the index date.

All measures were reported as descriptive statistics by group defined by influenza hospitalization status. Statistical differences between the groups were assessed using a two-sided t-test comparing means of continuous variables and Pearson’s chi-square test for categorical variables. A logistic regression model was used to estimate odds ratios (ORs) and 95% CIs for the association between patient demographic and baseline clinical characteristics and the odds of influenza-related hospitalization.

#### Stage 2

Patient demographic and baseline clinical characteristics were assessed during the 12-month baseline period. Follow-up measures were evaluated in the matched cohorts, including all-cause hospitalizations and emergency department (ED) visits within 30 days after the index date and during the index influenza season.

Baseline patient characteristics between the influenza and non-influenza cohorts before and after PS matching were assessed using standardized difference, applying a commonly used threshold of >10% in absolute standardized difference to determine imbalance [25]. Follow-up measures were compared using McNemar’s Chi-squared test for paired data. P-values <0.05 were considered statistically significant. Analyses were conducted using SAS version 9.4 (SAS Institute, Inc, Cary, NC).

## 3. Results

### 3.1. Stage 1 Results

#### 3.1.1 Study population

In total, 1,601,367 medically-attended influenza patients met the study inclusion criteria (**Fig. 1**). Of these, 18,509 (1.2%) patients had ≥1 influenza-related hospitalization within 30 days after the index date and 1,582,858 (98.8%) patients did not have evidence of hospitalization.

**Fig. 1.**
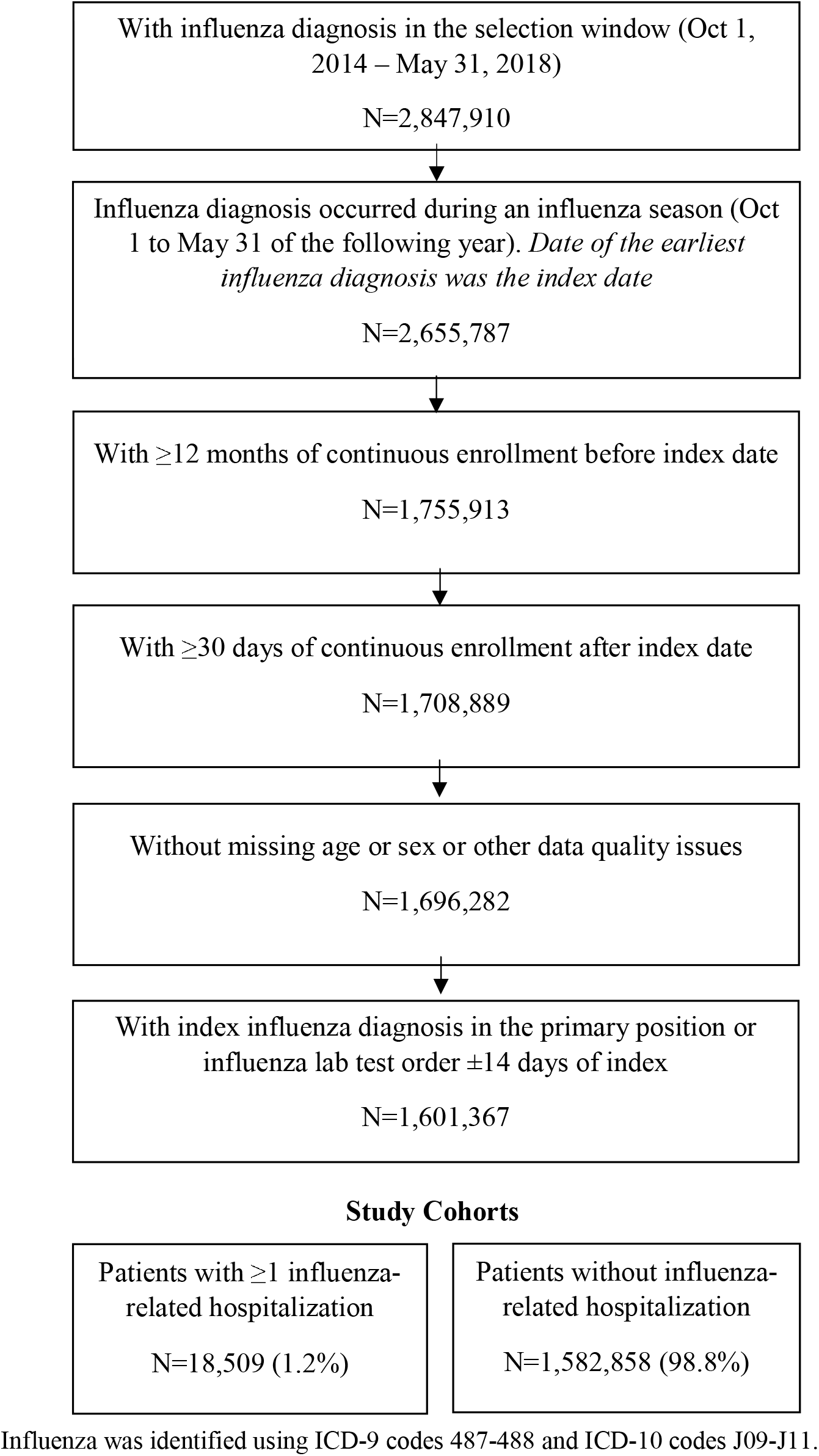
Stage 1 patient selection criteria Influenza was identified using ICD-9 codes 487-488 and ICD-10 codes J09-J11.

#### 3.1.2 Patient demographic and baseline characteristics

Medically-attended influenza patients who were hospitalized generally differed from those not hospitalized, in terms of baseline demographic and clinical characteristics (**Table 1**. Hospitalized patients were, on average, nearly 20 years older compared to non-hospitalized patients (mean age, 47.3 vs. 28.8 years; p<0.0001) and were more likely to be ≥65 years of age (20.5% vs. 2.3%). In addition, a larger proportion of hospitalized patients had Medicaid (9.6% vs. 6.9%) or Medicare Risk (4.6% vs. 0.2%) plans compared to non-hospitalized patients (p<0.0001). Hospitalized patients also had a higher baseline comorbidity burden (mean AHRQ/CDC comorbidities, 3.8 vs. 1.0, p<0.0001) and 43.6% of hospitalized patients had 4+ comorbidities compared to 7.7% of non-hospitalized patients (**Fig. 2**). Furthermore, in the entire sample of influenza patients, 5.8% of patients ages 65-74 and 25.7% of patients ages ≥75 were hospitalized within 30 days after the index date, compared to 2.3%, 0.8%, 0.4%, and 0.9% of patients ages 50-64, 18-49, 5-17, and 0-4, respectively. These age trends persisted among patients without evidence of any AHRQ/CDC comorbidities during baseline (**Fig. 3**). Lastly, 21.4% of hospitalized patients had evidence of an influenza vaccine during the respective influenza season in the claims database compared to 17.7% of non-hospitalized patients (p<0.0001).

**Table 1.** Baseline demographic and clinical characteristics by hospitalization status

| Baseline characteristics | Patients with $\geq 1$ influenza-related hospitalization<br>N = 18,509 | Patients without influenza-related hospitalization<br>N = 1,582,858 | p-value |
| --- | --- | --- | --- |
| **Age, mean±SD** | 47.3±23.1 | 28.8±19.8 | <0.0001 |
| **Age group, n (%)** |  |  | <0.0001 |
| 0-4 | 1,201 (6.5) | 125,906 (8.0) |  |
| 5-17 | 1,807 (9.8) | 512,517 (32.4) |  |
| 18-49 | 4,926 (26.6) | 623,281 (39.4) |  |
| 50-64 | 6,787 (36.7) | 284,492 (18.0) |  |
| 65+ | 3,788 (20.5) | 36,662 (2.3) |  |
| **Sex, n (%)** |  |  | 0.71 |
| Male | 8,749 (47.3) | 745,993 (47.1) |  |
| Female | 9,760 (52.7) | 836,865 (52.0) |  |
| **Payer type, n (%)** |  |  | <0.0001 |
| Commercial | 9,374 (50.7) | 866,656 (54.8) |  |
| Self-insured | 6,426 (34.7) | 599,540 (37.9) |  |
| Medicaid | 1,769 (9.6) | 108,617 (6.9) |  |
| Medicare risk | 852 (4.6) | 2,509 (0.2) |  |
| Other/Unknown | 88 (0.5) | 5,536 (0.3) |  |
| **Geographic region, n (%)** |  |  | <0.0001 |
| Northeast | 3,457 (18.7) | 188,490 (11.9) |  |
| Midwest | 4,758 (25.7) | 334,387 (21.1) |  |
| South | 7,583 (41.0) | 906,543 (57.3) |  |
| West | 2,711 (14.7) | 153,438 (9.7) |  |
| **AHRQ/CDC comorbidities\*, mean±SD** | 3.8±3.5 | 1.0±1.6 | <0.0001 |
| **AHRQ/CDC comorbidities category\*, n (%)** |  |  | <0.0001 |
| 0 | 3,510 (19.0) | 891,422 (56.3) |  |
| 1-3 | 6,924 (37.4) | 569,667 (36.0) |  |
| 4-6 | 4,456 (24.1) | 102,322 (6.5) |  |
| 7-9 | 2,221 (12.0) | 15,774 (1.0) |  |
| 10+ | 1,396 (7.5) | 3,421 (0.2) |  |
| **Index influenza season, n (%)** |  |  | <0.0001 |
| 2014 | 4,725 (25.5) | 444,002 (28.1) |  |
| 2015 | 3,533 (19.1) | 230,985 (14.6) |  |
| 2016 | 4,373 (23.6) | 379,894 (24.0) |  |
| 2017 | 5,878 (31.8) | 527,977 (33.4) |  |
| **Evidence of influenza vaccination before influenza diagnosis in the index season\*,**<br>**n (%)** | 3,954 (21.4) | 280,898 (17.7) | <0.0001 |

AHRQ: Agency for Health Research and Quality, CDC: Centers for Disease Control and Prevention, SD: standard deviation, SD: standard deviation

**Fig. 2.**
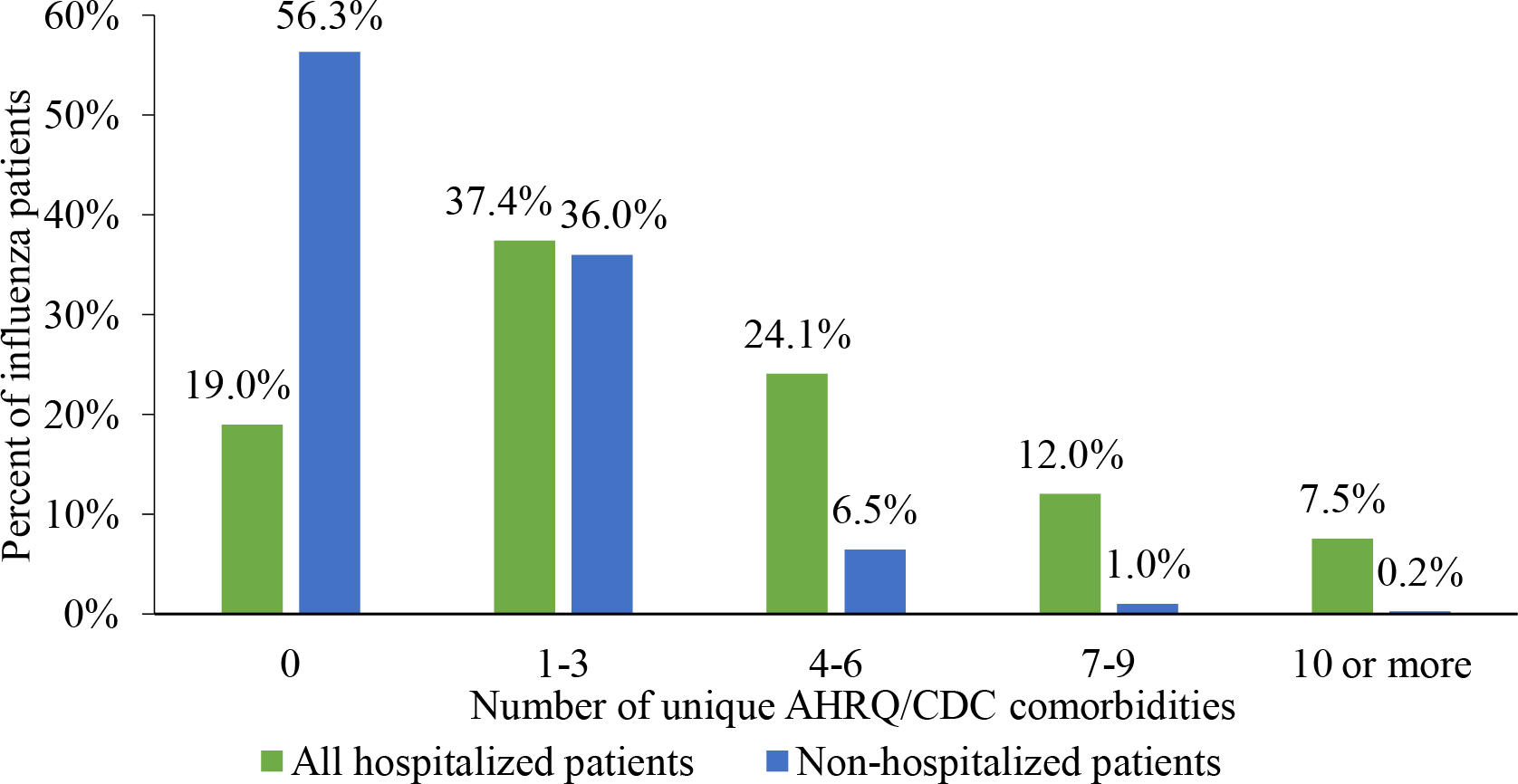
Number of baseline AHRQ/CDC comorbidities by influenza-related hospitalization status AHRQ: Agency for Health Research and Quality, CDC: Centers for Disease Control and Prevention *Patients could have a maximum of 37 comorbidities defined using ICD-9/ICD-10 codes from the Elixhauser Comorbidity Software (AHRQ) and groups defined by the CDC as at high-risk for influenza-related hospitalizations.

**Fig. 3.**
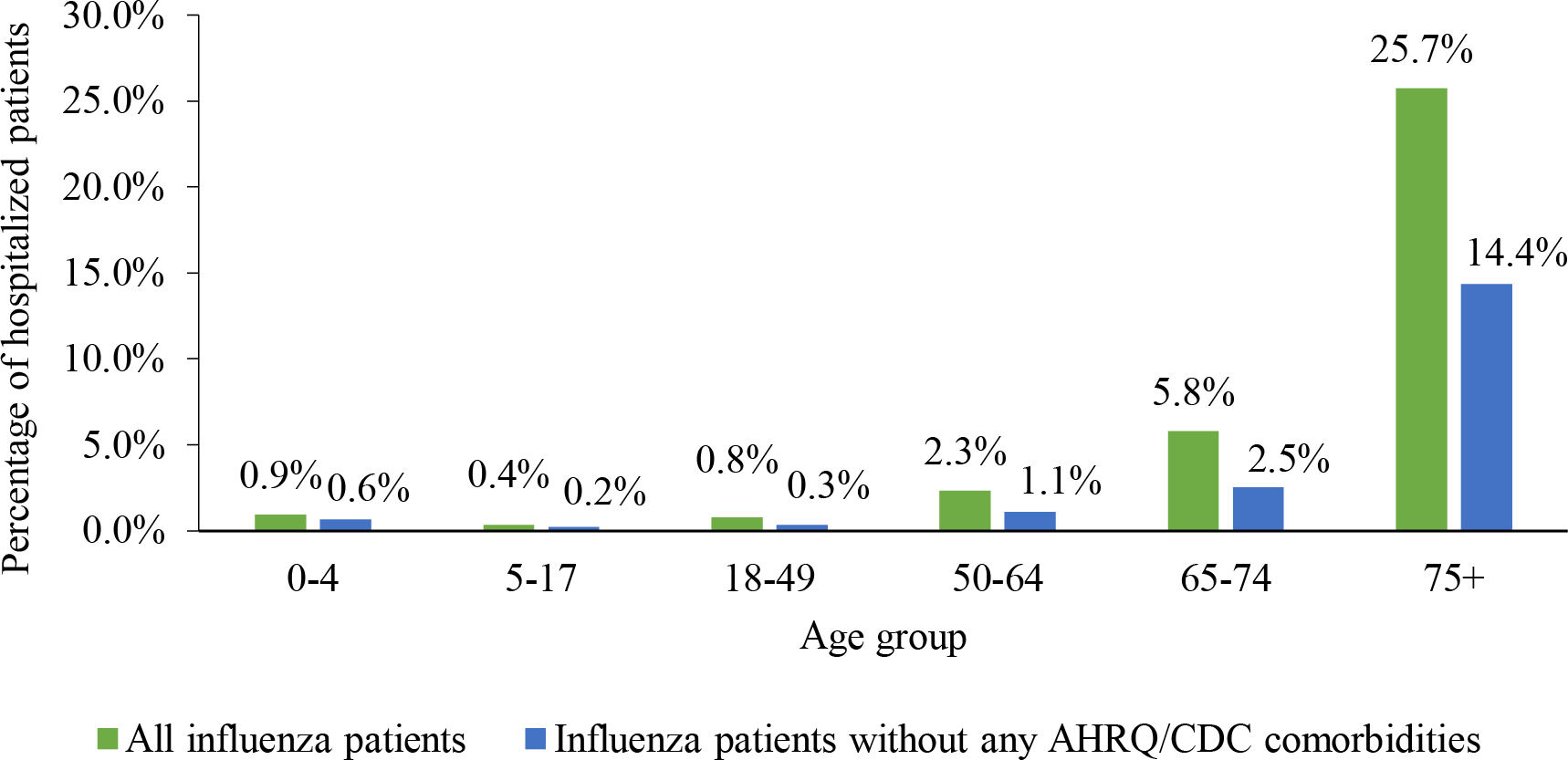
Influenza-related hospitalization rates of patients without any AHRQ/CDC comorbidities in the baseline period compared to all influenza patients AHRQ: Agency for Health Research and Quality, CDC: Centers for Disease Control and Prevention

#### 3.1.3 Predictors of influenza-related hospitalization

Elderly age and specific comorbidities were identified as predictors of influenza-related hospitalization in the logistic regression model, adjusted for baseline demographic and clinical characteristics (**Fig. 4**). Specifically, elderly patients (≥65 years) had 9.4 times higher odds of hospitalization compared to patients aged 5-17 years (95% CI, 8.8-10.1). Patients with certain groupings of comorbidities during baseline also had 2-3 times the odds of influenza-related hospitalization compared to patients without those underlying conditions, including leukemia, lymphoma and/or metastatic cancer; neuromuscular disease and/or paralysis, immunodeficiency and/or Human Immunodeficiency Virus/Acquired Immunodeficiency Syndrome [HIV/AIDS]; and COPD and/or chronic pulmonary disease) (all p<0.0001).

**Fig. 4.**
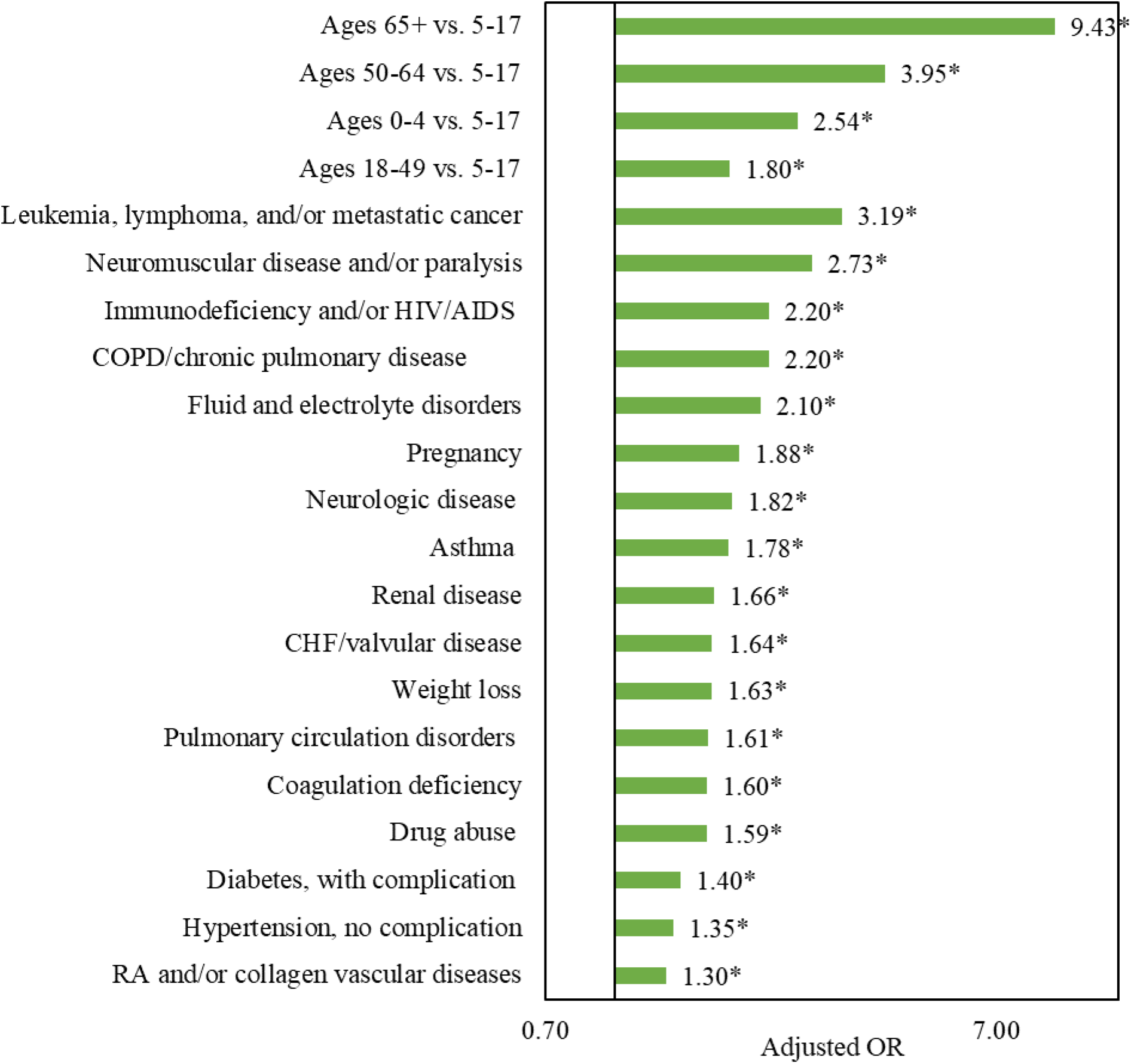
Logistic regression model for the odds of influenza-related hospitalization, adjusting for baseline demographic and clinical characteristics CHF: Congestive heart failure, COPD: Chronic obstructive pulmonary disease, HIV/AIDS: Human Immunodeficiency Virus/Acquired Immunodeficiency Syndrome *Indicates significant difference between hospitalized and non-hospitalized patients (p<0.05). Logistic regression model adjusted for age, sex, payer type, geographic region, index influenza season, evidence of influenza vaccination, and specific AHRQ/CDC comorbidities. This figure shows the odds ratios only for age groups and for select comorbidities where the odds ratio was >1.30. Other comorbidities significantly associated with the odds of influenza-related hospitalization included rheumatoid arthritis/collagen vascular diseases, diabetes with no complication, deficiency anemias, solid tumor without metastasis, alcohol abuse, hypertension with complication, peripheral vascular disorder, liver disease, psychoses, and depression (odds ratios ranging from 1.08 to 1.30). Blood loss anemia, obesity, other metabolic disease, hypothyroidism, and chronic peptic ulcer disease were not associated with the odds of influenza-related hospitalization.

### 3.2. Stage 2 Results

#### 3.2.1 Study population

Given the higher odds of influenza-related hospitalization observed among elderly patients in Stage 1, we identified a sample of influenza and non-influenza patients ages ≥65 years from October 1, 2013 – March 31, 2019. In total, 46,263 influenza and 1,103,034 non-influenza patients met study selection criteria (**Fig. 5**) and were further categorized into 12 influenza and 12 non-influenza cohorts with ≥1 comorbidity of interest. Patient demographic and baseline clinical characteristics and baseline total all-cause healthcare costs were well-balanced after 1:1 PS-matching. Cohort sizes and characteristics after PS-matching are provided in **Supplementary Tables 2 and 3**.

**Fig. 5.**
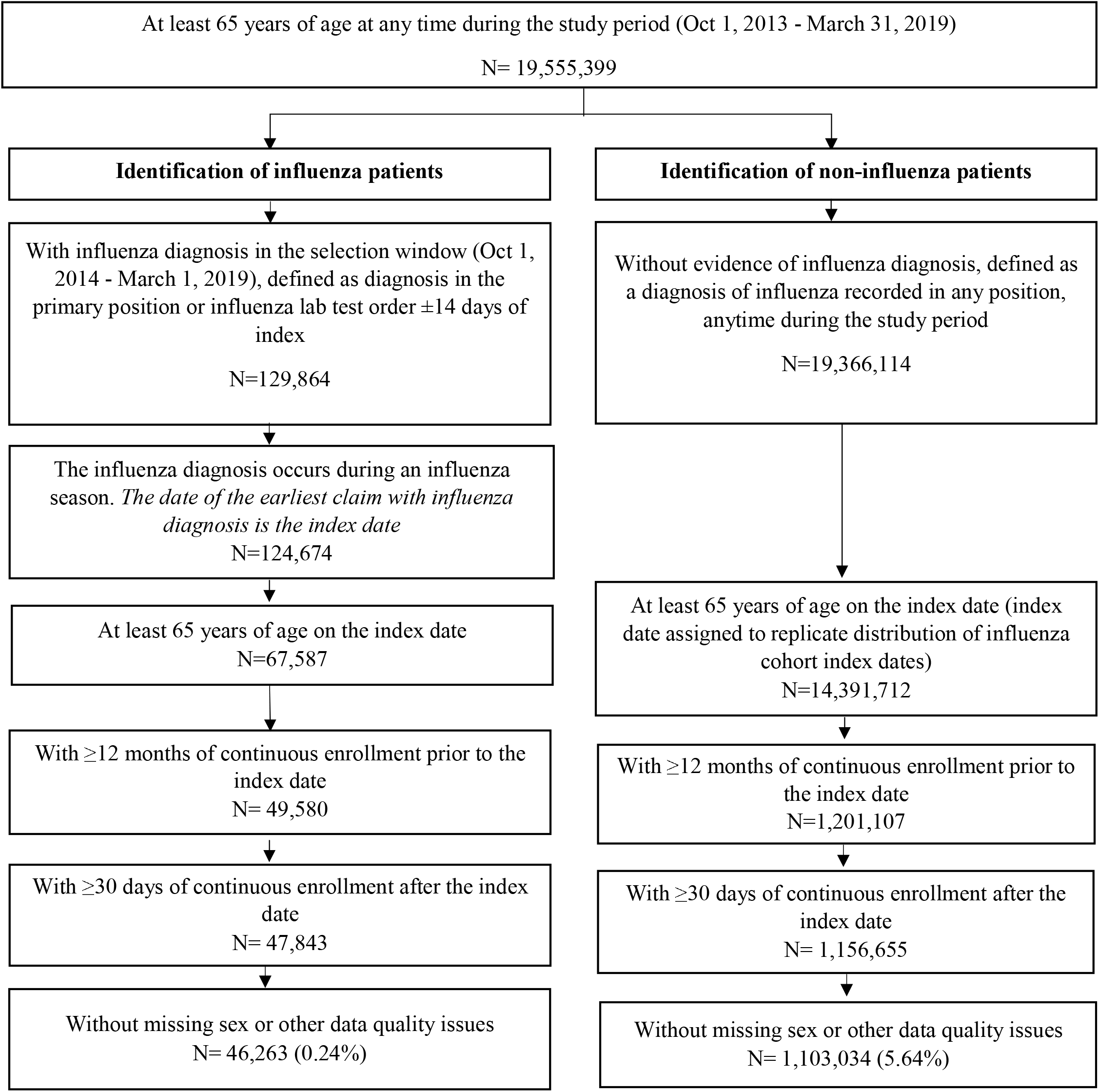
Stage 2 patient selection criteria

#### 3.2.1 All-cause hospitalization and ED visits during follow-up

Overall, the frequency of ≥1 all-cause hospitalization was higher for the influenza cohort during the 30-day follow-up period and during the index influenza season compared to matched controls for all comorbidity subgroups. During the 30-day follow-up period, the influenza cohorts had 3-7 times higher rates of hospitalization compared to the corresponding non-influenza cohorts; these differences were most prominent for CHF (41.0% vs. 7.9%), COPD (34.6% vs. 6.1%), CAD (22.8% vs. 3.8%), and late stage CKD (44.1% vs. 13.1%; all p<0.0001) (**Fig. 6a**). The differences were smaller when broadening follow-up to the influenza season, with 2-3 times higher hospitalization rates for the influenza cohorts compared to the matched non-influenza cohorts: CHF (48.2% vs. 20.7%), COPD (40.5% vs. 15.3%), CAD (27.8% vs. 10.8%), or late stage CKD (53.6% vs. 25.9%; all p<0.0001) (**Fig. 6b**). In Stage 1, the mean cost of influenza-related hospitalization among the hospitalized cohort was $22,169 (standard deviation [SD], $61,593) and the median cost was $11,014.8 (quartile 1, $6,653; quartile 3; $19,246).

**Fig. 6.**
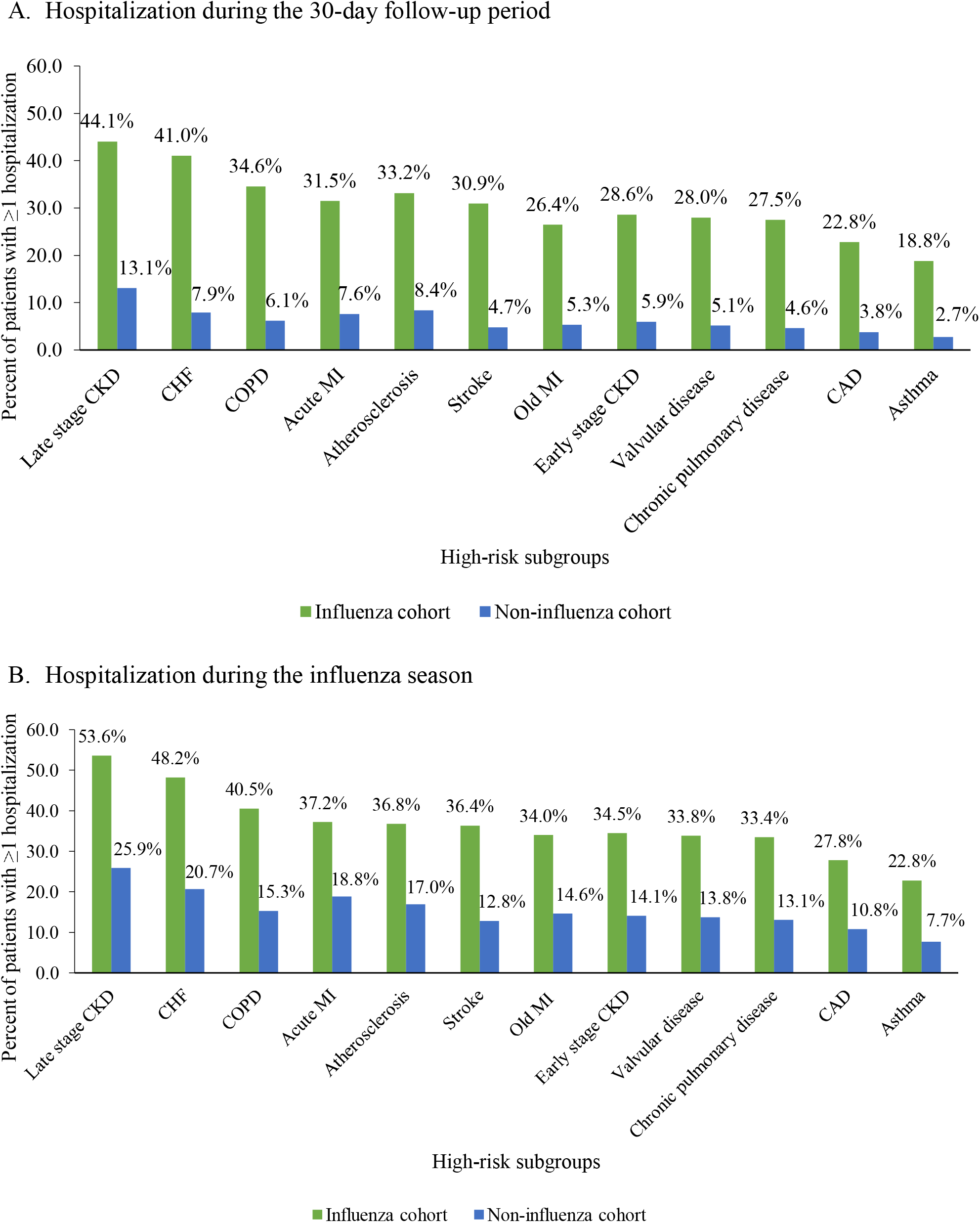
All-cause hospitalization rates in the elderly influenza and non-influenza cohorts by comorbidity status CAD: Coronary artery disease, CHF: Congestive heart failure, CKD: Chronic kidney disease, COPD: Chronic obstructive pulmonary disease, MI: myocardial infarction All p-values for comparisons of hospitalization rates between the influenza and non-influenza cohorts <0.05.

Similar findings were also observed for ED visits. During the 30-day follow-up period, influenza patients with CHF, COPD, CAD, or late stage CKD had 4-6 times higher ED visit rates compared to the corresponding non-influenza cohort (37.2% vs. 9.3%, 34.9% vs. 6.2%, 29.4% vs. 5.2%, 45.2% vs. 10.3%, respectively; all p<0.0001) **(Fig. 7a**). During the influenza season, ED visit rates were about two times higher in influenza patients compared to non-influenza patients; all p<0.0001) (**Fig. 7b**).

**Fig. 7.**
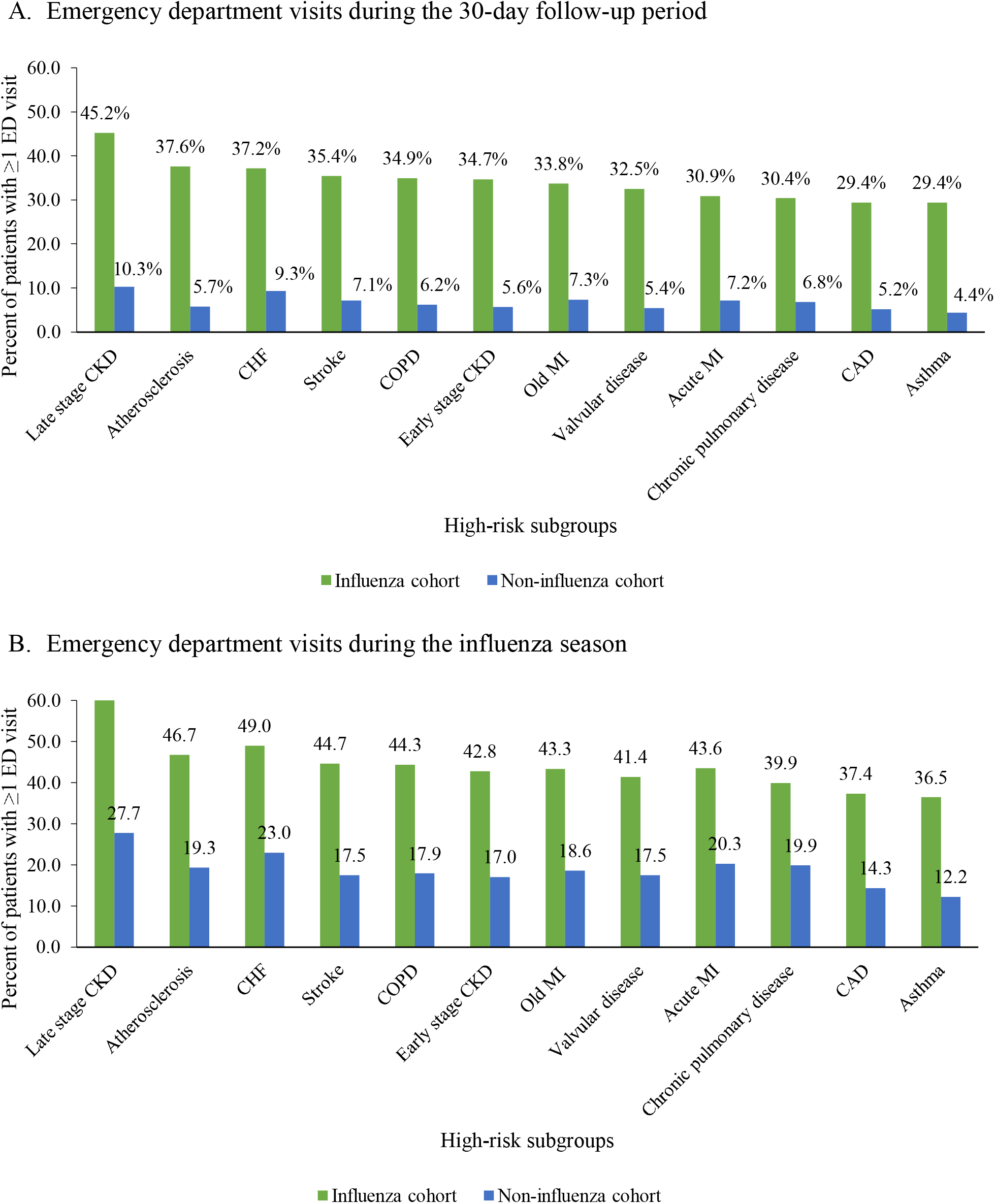
Emergency department visits during the 30-day follow-up period in the elderly influenza and non-influenza cohorts by comorbidity status CAD: Coronary artery disease, CHF: Congestive heart failure, CKD: Chronic kidney disease, COPD: Chronic obstructive pulmonary disease, MI: myocardial infarction All p-values for comparisons of ED visit rates between the influenza and non-influenza cohorts <0.05.

## 4. Discussion

The findings of this real-world study demonstrate that the elderly population and those with specific comorbidities are at elevated risk of influenza-related complications that may result in hospitalizations, a primary driver of the substantial economic burden of the disease. While the severity of influenza can vary with every season, the susceptibility of the elderly to disease-related complications has been consistently reported in epidemiological studies across geographies and has been attributed to alterations in immune defenses with age [26]. In our study, we identified more than 1.6 million influenza patients. Of these patients, only 2.5% were elderly and yet they accounted for 20.5% of influenza-related hospitalizations. Apart from age-related vulnerability, our study also identified that certain pre-existing comorbidities have a direct impact on outcomes, increasing the odds of hospitalization by 2 to 3-fold. Similar findings were reported in a meta-analysis of populations at risk for severe or complicated seasonal influenza, which found that the elderly had higher risk of hospitalization and death compared to the non-elderly (OR, 3.0; 95% CI, 1.5-5.7). Additionally, comorbidities were strongly associated with death (OR, 2.0; 95% CI, 1.7-2.4) and conditions like chronic lung disease, cardiovascular disease, COPD, and diabetes increased the probability of hospitalization and, in some cases, ventilator support [10].

Seasonal influenza-related complications and hospitalizations are not only life-threatening to populations at risk, but also a substantial economic burden for the healthcare system [2] and persistent efforts are needed to alleviate the burden in the high-risk populations highlighted in this study through vaccinations, early treatment, and existing prophylactic medications [21] or novel targeted therapies, such as monoclonal antibodies [27]. This strategy applies to the management of COVID-19 as well, since elderly age and comorbidities, including COPD, have also been implicated as risk factors for severe COVID-19 [28]. Although preliminary data suggest historically low influenza activity in the 2020-21 season due to the impact of COVID-19 and associated community mitigation measures (e.g., social distancing and mask wearing), preparation for future influenza seasons and potential co-epidemics of influenza and COVID-19 is needed, if COVID-19 becomes a recurrent seasonal disease [29].

Currently, vaccination is the most promising method to prevent and control influenza by reducing the risk of ailment by 40-60% and reducing disease severity among patients with post-vaccination infections [30]. Although the US vaccination target of 70% [31] was not achieved in the 2019-20 influenza season, with only 51.8% of the population aged ≥6 months receiving a vaccination [32], the impact of vaccination on morbidity and mortality was sizeable. Using a model accounting for vaccination coverage, vaccine efficacy, and disease occurrence, the CDC estimated that vaccinations prevented 7.5 million influenza illnesses, 3.7 million influenza-associated medical visits, 105,000 influenza-associated hospitalizations, and 6,300 influenza-associated deaths in the 2019-20 season [33, 34]. Similarly, despite observed “breakthrough” cases of COVID-19, the CDC recently reported that the risks of hospitalization and death were more than ten times lower among fully vaccinated vs. not fully vaccinated individuals in the US from April to July 2021 [35] and another study by Gupta et al. estimated that the US COVID-19 vaccination campaign prevented up to 140,000 deaths from December 2020 to May 2021 [36].

Achieving the vaccination targets for influenza or COVID-19 would allow for even larger impacts, but complacency and concerns regarding safety and efficacy are factors responsible for lack of widespread vaccination [37, 38]. For the elderly population at risk for complications, the vaccination target in the US is even higher at 90%, yet this target is rarely reached in the industrialized world [39]. However, the effectiveness of the vaccine in the elderly is known to be reduced due to lower seroconversion rates from poorer immunologic response [40] and in Stage 1 of this study, we observed higher rates of vaccination among influenza patients with hospitalization (21.4%) compared to patients without hospitalization (17.7%). Hence, despite the efforts towards widespread vaccination programs, there remains an unmet need for preventing complications of influenza in vulnerable populations, even among those who received vaccinations.

The strength of our study lies in estimating influenza-related hospitalizations in the backdrop of risk factors like pre-existing comorbidities, thus adding to the current literature related to influenza-related HRU using the most recently available data at the time of the study. The existing CDC model for reporting influenza-associated hospitalizations uses the reported number of hospitalizations to calculate hospitalization rates, which are adjusted to compensate for any under-detection. The adjusted rates are then applied to the US population by age group to estimate total number of influenza-related hospitalizations [41]. This method is limited due to lack of inclusion of the full denominator of patient groups at-risk for influenza because of underlying comorbid conditions, which can result in inadequacies when assessing the actual clinical and economic burden.

Several limitations of the retrospective nature of this analysis must be noted. First, clinical data were not available in this study, which may have led to misclassification of the influenza diagnoses due to the inability to verify disease status with test results. To account for this, we required evidence of an influenza lab test within 14 days of influenza diagnosis (in any position) or primary diagnosis of influenza on the claim. This study likely also underestimates the receipt of influenza vaccinations prior to the index date since we observed only about one-third of patients across the influenza and non-influenza cohorts with documented vaccination in the claims database. Only vaccinations documented by insurance would be captured in the study database; free vaccines or vaccines paid by cash are not captured. In addition, we did not investigate outcomes like rehabilitation and nursing home care after hospitalization, which contribute to the long-term disease burden not captured in the present study and should be evaluated in future real-world studies using longer follow-up periods. Lastly, given the study population was restricted to the commercially insured population, our findings may not be generalizable to the elderly population insured through traditional Medicare. This may be reflected in the relatively lower proportion of hospitalized influenza patients in our study ≥65 years (20.5%) compared to statistics reported by the CDC (50-70%) [26].

Overall, this real-world study reinforces previous research demonstrating a high clinical and economic burden attributable to influenza in elderly populations, but by varying degrees depending on their baseline comorbidity profile. Continued efforts to reduce influenza burden through prophylaxis are needed in high-risk populations in the US and globally.

## Data Availability

All data produced in the present work are contained in the manuscript

## ACKNOWLEDGEMENTS

The authors would like to acknowledge the contributions of Dr. Xiao Ding of VIR Biotechnology for the statistical review of the protocol, analysis plan, and results, Hsiu-Ching Chang of IQVIA for conducting the programming and statistical analysis, and Kasturi Chatterjee for manuscript writing assistance.

## FUNDING

This work was supported by VIR Biotechnology Inc.

## CONFLICT OF INTEREST STATEMENT

D.H. and C.R. report financial support from VIR Biotechnology Inc. Y.Y. reports a relationship with VIR Biotechnology Inc. that includes: funding grants. D.H. reports a relationship with Janssen Pharmaceuticals Inc. that includes: employment. D.H. reports a relationship with VIR Biotechnology Inc. that includes: equity or stocks. C.R. reports a relationship with VIR Biotechnology Inc that includes: employment and equity or stocks. A.N. and J.T. are employees of IQVIA, which received funding from VIR Biotechnology Inc. to conduct this study.

## Notes

### Competing Interest Statement

Carolina Reyes is employed by VIR Biotechnology Inc. David Hong was employed by VIR Biotechnology Inc at the time of this study. Yinong Young-Xu has received funding grants from VIR Biotechnology Inc. Aimee Near and Jenny Tse are employed by IQVIA, which received funding from VIR Biotechnology Inc to conduct this study.

### Funding Statement

This study was funded by VIR Biotechnology Inc.

### Summary of Updates

The online version of the manuscript has no values in Figure 4. They appear as [VALUE]. A non-editable version of Figure 4 has been provided in the attached manuscript. No changes to Supplement.

