## Supplemental File for "Burden of influenza hospitalization among high-risk groups in the United States"

**Supplementary data**

**Abbreviations**

| CAD | Coronary artery disease |
| --- | --- |
| CCI | Charlson comorbidity index |
| CHF | Congestive heart failure |
| CKD | Chronic kidney disease |
| COPD | Chronic obstructive pulmonary disease |
| HMO | Health maintenance organization |
| MI | Myocardial infarction |
| POS | Point of service |
| PPO | Preferred provider organization |
| PS | Propensity score |
| SD | Standard deviation |
| Std. Diff. | Standardized difference |

### Supplementary Table 1. Samples sizes of the influenza and non-influenza cohorts before and after propensity score matching

| Study Populations (All Patients Ages 65+ in Index Year) | Sample Size before PS-Matching | | Total Size of 1:1 PS-Matched Sample* |
| --- | --- | --- | --- |
|  | **Influenza cohort** | **Non-Influenza cohort** |  |
| Asthma | 2,202 | 23,194 | 4,400 |
| COPD | 2,767 | 36,984 | 5,534 |
| Chronic pulmonary disease | 590 | 5,568 | 1,178 |
| Atherosclerosis | 386 | 5,022 | 766 |
| CAD | 5,112 | 80,761 | 10,222 |
| CHF | 1,931 | 25,646 | 3,860 |
| Stroke | 1,288 | 20,683 | 2,574 |
| Valvular disease | 1,380 | 23,048 | 2,760 |
| Old MI† | 398 | 5,677 | 794 |
| Acute MI† | 477 | 6,663 | 946 |
| Early stage CKD^‡^ | 2,021 | 31,763 | 4,040 |
| Late stage CKD^‡^ | 432 | 5,601 | 858 |

* Factors used for propensity score matching were age category (exact match), sex (exact match), payer type, plan type, geographic region, Charlson comorbidity index category, specific comorbidities, evidence of influenza vaccine in the index influenza season before the index date, index influenza season, index month, and baseline total healthcare costs (log-transformed, continuous)

† Presence of diagnosis code for acute MI was prioritized over old MI; old MI and acute MI are mutually exclusive

^‡^ Presence of diagnosis code for late stage CKD was prioritized over early stage CKD; these severity groups are mutually exclusive

### Supplementary Table 2. Baseline demographic clinical characteristics for influenza and non-influenza patients with baseline asthma after PS-matching

|  | Asthma | | | | |
| --- | --- | --- | --- | --- | --- |
| Measures* | Influenza cohort | | Non-influenza cohort | | Std. Diff. |
|  | N=2,200 | | N=2,200 | |  |
|  | N | % | N | % |  |
| Age categories, years |  |  |  |  |  |
| 65-74 | 1,722 | 78.3 | 1,722 | 78.3 | 0.00 |
| 75+ | 478 | 21.7 | 478 | 21.7 |  |
| Sex |  |  |  |  |  |
| Male | 743 | 33.8 | 743 | 33.8 | 0.00 |
| Female | 1,457 | 66.2 | 1,457 | 66.2 |  |
| Geographic region |  |  |  |  |  |
| Northeast | 516 | 23.5 | 497 | 22.6 | 0.02 |
| Midwest | 388 | 17.6 | 388 | 17.6 |  |
| South | 792 | 36.0 | 793 | 36.0 |  |
| West | 504 | 22.9 | 522 | 23.7 |  |
| Payer type |  |  |  |  |  |
| Commercial | 1,084 | 49.3 | 1,102 | 50.1 | 0.05 |
| Self-insured | 774 | 35.2 | 767 | 34.9 |  |
| Medicaid | 72 | 3.3 | 76 | 3.5 |  |
| Medicare risk | 248 | 11.3 | 242 | 11.0 |  |
| Other/Unknown | 22 | 1.0 | 13 | 0.6 |  |
| Plan type |  |  |  |  |  |
| HMO | 357 | 16.2 | 339 | 15.4 | 0.07 |
| PPO | 1,685 | 76.6 | 1,732 | 78.7 |  |
| POS | 51 | 2.3 | 34 | 1.5 |  |
| Indemnity | 77 | 3.5 | 73 | 3.3 |  |
| Other/Unknown | 30 | 1.4 | 22 | 1.0 |  |
| Charlson comorbidity index categories |  |  |  |  |  |
| 0 | 0 | 0 | 0 | 0 | 0.04 |
| 1 | 787 | 35.8 | 818 | 37.2 |  |
| 2 | 426 | 19.4 | 440 | 20.0 |  |
| 3 | 347 | 15.8 | 321 | 14.6 |  |
| ≥4 | 640 | 29.1 | 621 | 28.2 |  |
| Specific comorbidities |  |  |  |  |  |
| COPD | 430 | 19.5 | 435 | 19.8 | 0.01 |
| Chronic pulmonary disease | 137 | 6.2 | 117 | 5.3 | -0.04 |
| Asthma | 2,200 | 100.0 | 2,200 | 100.0 | 0.00 |
| CHF | 203 | 9.2 | 213 | 9.7 | 0.02 |
| Valvular disease | 132 | 6.0 | 118 | 5.4 | -0.03 |
| CAD | 317 | 14.4 | 294 | 13.4 | -0.03 |
| Old MI | 27 | 1.2 | 19 | 0.9 | -0.04 |
| Acute MI | 49 | 2.2 | 54 | 2.5 | 0.02 |
| Stroke | 102 | 4.6 | 83 | 3.8 | -0.04 |
| Atherosclerosis | 30 | 1.4 | 28 | 1.3 | -0.01 |
| Early stage CKD | 176 | 8.0 | 177 | 8.0 | 0.002 |
| Late stage CKD | 27 | 1.2 | 26 | 1.2 | -0.004 |
| Influenza vaccination in the index influenza season before the index date | 729 | 33.1 | 727 | 33.0 | -0.002 |
| Index influenza season |  |  |  |  |  |
| 2014 (Oct 2014 - May 2015) | 570 | 25.9 | 595 | 27.0 | 0.03 |
| 2015 (Oct 2015 - May 2016) | 235 | 10.7 | 225 | 10.2 |  |
| 2016 (Oct 2016 - May 2017) | 540 | 24.5 | 524 | 23.8 |  |
| 2017 (Oct 2017 - May 2018) | 660 | 30.0 | 670 | 30.5 |  |
| 2018 (Oct 2018 - March 2019) | 195 | 8.9 | 186 | 8.5 |  |
| Index month |  |  |  |  |  |
| October | 42 | 1.9 | 41 | 1.9 | 0.05 |
| November | 50 | 2.3 | 43 | 2.0 |  |
| December | 320 | 14.5 | 324 | 14.7 |  |
| January | 686 | 31.2 | 693 | 31.5 |  |
| February | 601 | 27.3 | 630 | 28.6 |  |
| March | 323 | 14.7 | 299 | 13.6 |  |
| April | 132 | 6.0 | 125 | 5.7 |  |
| May | 46 | 2.1 | 45 | 2.0 |  |
| Total all-cause healthcare costs during the 12-month baseline period ($) |  |  |  |  |  |
| Mean (SD) | $39,021 | $102,820 | $37,193 | $69,834 | 0.001 |
| * Demographic measures (age, sex, payer type, plan type, and geographic region) and timing of index date (index influenza season and index month) were reported from data on the index date. Baseline clinical characteristics (Charlson comorbidity index, specific comorbidities, evidence of influenza vaccine, and baseline total healthcare costs) were reported using data from the 12-month baseline period. | | | | | |

### Supplementary Table 3. Baseline demographic and clinical characteristics for influenza and non-influenza patients with baseline COPD after PS-matching

|  | COPD | | | | |
| --- | --- | --- | --- | --- | --- |
| Measures* | Influenza cohort | | Non-influenza cohort | | Std. Diff. |
|  | N=2,767 | | N=2,767 | |  |
|  | N | % | N | % |  |
| Age categories, years |  |  |  |  |  |
| 65-74 | 1,771 | 64.0 | 1,771 | 64.0 | 0.00 |
| 75+ | 996 | 36.0 | 996 | 36.0 |  |
| Sex |  |  |  |  |  |
| Male | 1,395 | 50.4 | 1,395 | 50.4 | 0.00 |
| Female | 1,372 | 49.6 | 1,372 | 49.6 |  |
| Geographic region |  |  |  |  |  |
| Northeast | 689 | 24.9 | 736 | 26.6 | 0.05 |
| Midwest | 518 | 18.7 | 520 | 18.8 |  |
| South | 1,010 | 36.5 | 1,009 | 36.5 |  |
| West | 550 | 19.9 | 502 | 18.1 |  |
| Payer type |  |  |  |  |  |
| Commercial | 1,198 | 43.3 | 1,201 | 43.4 | 0.02 |
| Self-insured | 965 | 34.9 | 949 | 34.3 |  |
| Medicaid | 112 | 4.0 | 112 | 4.0 |  |
| Medicare risk | 471 | 17.0 | 486 | 17.6 |  |
| Other/Unknown | 21 | 0.8 | 19 | 0.7 |  |
| Plan type |  |  |  |  |  |
| HMO | 509 | 18.4 | 504 | 18.2 | 0.01 |
| PPO | 2,053 | 74.2 | 2,059 | 74.4 |  |
| POS | 60 | 2.2 | 61 | 2.2 |  |
| Indemnity | 118 | 4.3 | 119 | 4.3 |  |
| Other/Unknown | 27 | 1.0 | 24 | 0.9 |  |
| Charlson comorbidity index categories |  |  |  |  |  |
| 0 | 0 | 0.0 | 0 | 0.0 | 0.03 |
| 1 | 598 | 21.6 | 581 | 21.0 |  |
| 2 | 435 | 15.7 | 434 | 15.7 |  |
| 3 | 482 | 17.4 | 464 | 16.8 |  |
| ≥4 | 1,252 | 45.2 | 1,288 | 46.5 |  |
| Specific comorbidities |  |  |  |  |  |
| COPD | 2,767 | 100.0 | 2,767 | 100.0 | 0.00 |
| Chronic pulmonary disease | 210 | 7.6 | 209 | 7.6 | 0.00 |
| Asthma | 432 | 15.6 | 438 | 15.8 | 0.01 |
| CHF | 601 | 21.7 | 643 | 23.2 | 0.04 |
| Valvular disease | 234 | 8.5 | 242 | 8.7 | 0.01 |
| CAD | 856 | 30.9 | 900 | 32.5 | 0.03 |
| Old MI | 102 | 3.7 | 98 | 3.5 | -0.01 |
| Acute MI | 123 | 4.4 | 130 | 4.7 | 0.01 |
| Stroke | 240 | 8.7 | 257 | 9.3 | 0.02 |
| Atherosclerosis | 94 | 3.4 | 100 | 3.6 | 0.01 |
| Early stage CKD | 327 | 11.8 | 360 | 13.0 | 0.04 |
| Late stage CKD | 99 | 3.6 | 118 | 4.3 | 0.04 |
| Influenza vaccination in the index influenza season before the index date | 852 | 30.8 | 842 | 30.4 | -0.01 |
| Index influenza season |  |  |  |  |  |
| 2014 (Oct 2014 - May 2015) | 785 | 28.4 | 800 | 28.9 | 0.04 |
| 2015 (Oct 2015 - May 2016) | 304 | 11.0 | 285 | 10.3 |  |
| 2016 (Oct 2016 - May 2017) | 653 | 23.6 | 661 | 23.9 |  |
| 2017 (Oct 2017 - May 2018) | 798 | 28.8 | 820 | 29.6 |  |
| 2018 (Oct 2018 - March 2019) | 227 | 8.2 | 201 | 7.3 |  |
| Index month |  |  |  |  |  |
| October | 44 | 1.6 | 44 | 1.6 | 0.06 |
| November | 76 | 2.7 | 74 | 2.7 |  |
| December | 444 | 16.0 | 482 | 17.4 |  |
| January | 868 | 31.4 | 863 | 31.2 |  |
| February | 752 | 27.2 | 708 | 25.6 |  |
| March | 366 | 13.2 | 378 | 13.7 |  |
| April | 168 | 6.1 | 179 | 6.5 |  |
| May | 49 | 1.8 | 39 | 1.4 |  |
| Total all-cause healthcare costs during the 12-month baseline period ($) |  |  |  |  |  |
| Mean (SD) | $54,924 | $134,582 | $56,359 | $125,697 | -0.0001 |
| * Demographic measures (age, sex, payer type, plan type, and geographic region) and timing of index date (index influenza season and index month) were reported from data on the index date. Baseline clinical characteristics (Charlson comorbidity index, specific comorbidities, evidence of influenza vaccine, and baseline total healthcare costs) were reported using data from the 12-month baseline period. | | | | | |

### Supplementary Table 4. Baseline demographic and clinical characteristics for influenza and non-influenza patients with baseline chronic pulmonary disease after PS-matching

|  | Chronic pulmonary disease | | | | |
| --- | --- | --- | --- | --- | --- |
| Measures* | Influenza cohort | | Non-influenza cohort | | Std. Diff. |
|  | N=589 | | N=589 | |  |
|  | N | % | N | % |  |
| Age categories, years |  |  |  |  |  |
| 65-74 | 390 | 66.2 | 390 | 66.2 | 0.00 |
| 75+ | 199 | 33.8 | 199 | 33.8 |  |
| Sex |  |  |  |  |  |
| Male | 282 | 47.9 | 282 | 47.9 | 0.00 |
| Female | 307 | 52.1 | 307 | 52.1 |  |
| Geographic region |  |  |  |  |  |
| Northeast | 110 | 18.7 | 102 | 17.3 | 0.09 |
| Midwest | 96 | 16.3 | 98 | 16.6 |  |
| South | 226 | 38.4 | 210 | 35.7 |  |
| West | 157 | 26.7 | 179 | 30.4 |  |
| Payer type |  |  |  |  |  |
| Commercial | 301 | 51.1 | 303 | 51.4 | 0.08 |
| Self-insured | 188 | 31.9 | 184 | 31.2 |  |
| Medicaid | 18 | 3.1 | 14 | 2.4 |  |
| Medicare risk | 79 | 13.4 | 87 | 14.8 |  |
| Other/Unknown | 3 | 0.5 | 1 | 0.2 |  |
| Plan type |  |  |  |  |  |
| HMO | 80 | 13.6 | 92 | 15.6 | 0.06 |
| PPO | 469 | 79.6 | 456 | 77.4 |  |
| POS | 6 | 1.0 | 7 | 1.2 |  |
| Indemnity | 29 | 4.9 | 29 | 4.9 |  |
| Other/Unknown | 5 | 0.8 | 5 | 0.8 |  |
| Charlson comorbidity index categories |  |  |  |  |  |
| 0 | 38 | 6.5 | 40 | 6.8 | 0.06 |
| 1 | 126 | 21.4 | 134 | 22.8 |  |
| 2 | 99 | 16.8 | 86 | 14.6 |  |
| 3 | 74 | 12.6 | 76 | 12.9 |  |
| ≥4 | 252 | 42.8 | 253 | 43.0 |  |
| Specific comorbidities |  |  |  |  |  |
| COPD | 209 | 35.5 | 200 | 34.0 | -0.03 |
| Chronic pulmonary disease | 589 | 100.0 | 589 | 100.0 | 0.00 |
| Asthma | 139 | 23.6 | 128 | 21.7 | -0.04 |
| CHF | 110 | 18.7 | 110 | 18.7 | 0.00 |
| Valvular disease | 48 | 8.1 | 51 | 8.7 | 0.02 |
| CAD | 174 | 29.5 | 164 | 27.8 | -0.04 |
| Old MI | 16 | 2.7 | 14 | 2.4 | -0.02 |
| Acute MI | 28 | 4.8 | 27 | 4.6 | -0.01 |
| Stroke | 40 | 6.8 | 38 | 6.5 | -0.01 |
| Atherosclerosis | 25 | 4.2 | 18 | 3.1 | -0.06 |
| Early stage CKD | 80 | 13.6 | 90 | 15.3 | 0.05 |
| Late stage CKD | 24 | 4.1 | 19 | 3.2 | -0.05 |
| Influenza vaccination in the index influenza season before the index date | 207 | 35.1 | 212 | 36.0 | 0.02 |
| Index influenza season |  |  |  |  |  |
| 2014 (Oct 2014 - May 2015) | 136 | 23.1 | 128 | 21.7 | 0.12 |
| 2015 (Oct 2015 - May 2016) | 61 | 10.4 | 69 | 11.7 |  |
| 2016 (Oct 2016 - May 2017) | 154 | 26.1 | 130 | 22.1 |  |
| 2017 (Oct 2017 - May 2018) | 189 | 32.1 | 207 | 35.1 |  |
| 2018 (Oct 2018 - March 2019) | 49 | 8.3 | 55 | 9.3 |  |
| Index month |  |  |  |  |  |
| October | 4 | 0.7 | 4 | 0.7 | 0.10 |
| November | 11 | 1.9 | 10 | 1.7 |  |
| December | 76 | 12.9 | 85 | 14.4 |  |
| January | 193 | 32.8 | 199 | 33.8 |  |
| February | 159 | 27.0 | 146 | 24.8 |  |
| March | 101 | 17.1 | 90 | 15.3 |  |
| April | 31 | 5.3 | 41 | 7.0 |  |
| May | 14 | 2.4 | 14 | 2.4 |  |
| Total all-cause healthcare costs during the 12-month baseline period ($) |  |  |  |  |  |
| Mean (SD) | $71,433 | $194,896 | $67,006 | $160,513 | -0.003 |
| * Demographic measures (age, sex, payer type, plan type, and geographic region) and timing of index date (index influenza season and index month) were reported from data on the index date. Baseline clinical characteristics (Charlson comorbidity index, specific comorbidities, evidence of influenza vaccine, and baseline total healthcare costs) were reported using data from the 12-month baseline period. | | | | | |

### Supplementary Table 5. Baseline demographic and clinical characteristics for influenza and non-influenza patients with baseline atherosclerosis after PS-matching

|  | Atherosclerosis | | | | |
| --- | --- | --- | --- | --- | --- |
| Measures* | Influenza cohort | | Non-influenza cohort | | Std. Diff. |
|  | N=383 | | N=383 | |  |
|  | N | % | N | % |  |
| Age categories, years |  |  |  |  |  |
| 65-74 | 184 | 48.0 | 184 | 48.0 | 0.00 |
| 75+ | 199 | 52.0 | 199 | 52.0 |  |
| Sex |  |  |  |  |  |
| Male | 193 | 50.4 | 193 | 50.4 | 0.00 |
| Female | 190 | 49.6 | 190 | 49.6 |  |
| Geographic region |  |  |  |  |  |
| Northeast | 134 | 35.0 | 146 | 38.1 | 0.09 |
| Midwest | 53 | 13.8 | 48 | 12.5 |  |
| South | 107 | 27.9 | 96 | 25.1 |  |
| West | 89 | 23.2 | 93 | 24.3 |  |
| Payer type |  |  |  |  |  |
| Commercial | 152 | 39.7 | 161 | 42.0 | 0.09 |
| Self-insured | 131 | 34.2 | 122 | 31.9 |  |
| Medicaid | 15 | 3.9 | 12 | 3.1 |  |
| Medicare risk | 82 | 21.4 | 83 | 21.7 |  |
| Other/Unknown | 3 | 0.8 | 5 | 1.3 |  |
| Plan type |  |  |  |  |  |
| HMO | 65 | 17.0 | 71 | 18.5 | 0.07 |
| PPO | 279 | 72.8 | 272 | 71.0 |  |
| POS | 6 | 1.6 | 7 | 1.8 |  |
| Indemnity | 30 | 7.8 | 28 | 7.3 |  |
| Other/Unknown | 3 | 0.8 | 5 | 1.3 |  |
| Charlson comorbidity index categories |  |  |  |  |  |
| 0 | 0 | 0.0 | 0 | 0.0 | 0.09 |
| 1 | 35 | 9.1 | 31 | 8.1 |  |
| 2 | 41 | 10.7 | 40 | 10.4 |  |
| 3 | 58 | 15.1 | 70 | 18.3 |  |
| ≥4 | 249 | 65.0 | 242 | 63.2 |  |
| Specific comorbidities |  |  |  |  |  |
| COPD | 92 | 24.0 | 90 | 23.5 | -0.01 |
| Chronic pulmonary disease | 23 | 6.0 | 23 | 6.0 | 0.00 |
| Asthma | 28 | 7.3 | 30 | 7.8 | 0.02 |
| CHF | 97 | 25.3 | 81 | 21.1 | -0.10 |
| Valvular disease | 55 | 14.4 | 62 | 16.2 | 0.05 |
| CAD | 166 | 43.3 | 153 | 39.9 | -0.07 |
| Old MI | 19 | 5.0 | 20 | 5.2 | 0.01 |
| Acute MI | 28 | 7.3 | 26 | 6.8 | -0.02 |
| Stroke | 63 | 16.4 | 69 | 18.0 | 0.04 |
| Atherosclerosis | 383 | 100.0 | 383 | 100.0 | 0.00 |
| Early stage CKD | 84 | 21.9 | 65 | 17.0 | -0.13 |
| Late stage CKD | 26 | 6.8 | 27 | 7.0 | 0.01 |
| Influenza vaccination in the index influenza season before the index date | 113 | 29.5 | 117 | 30.5 | 0.02 |
| Index influenza season |  |  |  |  |  |
| 2014 (Oct 2014 - May 2015) | 27 | 7.0 | 18 | 4.7 | 0.14 |
| 2015 (Oct 2015 - May 2016) | 33 | 8.6 | 35 | 9.1 |  |
| 2016 (Oct 2016 - May 2017) | 127 | 33.2 | 114 | 29.8 |  |
| 2017 (Oct 2017 - May 2018) | 159 | 41.5 | 178 | 46.5 |  |
| 2018 (Oct 2018 - March 2019) | 37 | 9.7 | 38 | 9.9 |  |
| Index month |  |  |  |  |  |
| October | 8 | 2.1 | 3 | 0.8 | 0.15 |
| November | 9 | 2.3 | 7 | 1.8 |  |
| December | 38 | 9.9 | 46 | 12.0 |  |
| January | 116 | 30.3 | 112 | 29.2 |  |
| February | 129 | 33.7 | 138 | 36.0 |  |
| March | 51 | 13.3 | 50 | 13.1 |  |
| April | 25 | 6.5 | 22 | 5.7 |  |
| May | 7 | 1.8 | 5 | 1.3 |  |
| Total all-cause healthcare costs during the 12-month baseline period ($) |  |  |  |  |  |
| Mean (SD) | $80,003 | $220,331 | $75,006 | $161,749 | -0.007 |
| * Demographic measures (age, sex, payer type, plan type, and geographic region) and timing of index date (index influenza season and index month) were reported from data on the index date. Baseline clinical characteristics (Charlson comorbidity index, specific comorbidities, evidence of influenza vaccine, and baseline total healthcare costs) were reported using data from the 12-month baseline period. | | | | | |

### Supplementary Table 6. Baseline demographic and clinical characteristics for influenza and non-influenza patients with baseline coronary artery disease (CAD) after PS-matching

|  | CAD | | | | |
| --- | --- | --- | --- | --- | --- |
| Measures* | Influenza cohort | | Non-influenza cohort | | Std. Diff. |
|  | N=5,111 | | N=5,111 | |  |
|  | N | % | N | % |  |
| Age categories, years |  |  |  |  |  |
| 65-74 | 3,283 | 64.2 | 3,283 | 64.2 | 0.00 |
| 75+ | 1,828 | 35.8 | 1,828 | 35.8 |  |
| Sex |  |  |  |  |  |
| Male | 3,599 | 70.4 | 3,599 | 70.4 | 0.00 |
| Female | 1,512 | 29.6 | 1,512 | 29.6 |  |
| Geographic region |  |  |  |  |  |
| Northeast | 1,195 | 23.4 | 1,197 | 23.4 | 0.02 |
| Midwest | 849 | 16.6 | 856 | 16.7 |  |
| South | 2,257 | 44.2 | 2,280 | 44.6 |  |
| West | 810 | 15.8 | 778 | 15.2 |  |
| Payer type |  |  |  |  |  |
| Commercial | 2,366 | 46.3 | 2,379 | 46.5 | 0.02 |
| Self-insured | 1,888 | 36.9 | 1,848 | 36.2 |  |
| Medicaid | 94 | 1.8 | 100 | 2.0 |  |
| Medicare risk | 716 | 14.0 | 735 | 14.4 |  |
| Other/Unknown | 47 | 0.9 | 49 | 1.0 |  |
| Plan type |  |  |  |  |  |
| HMO | 696 | 13.6 | 702 | 13.7 | 0.01 |
| PPO | 3,995 | 78.2 | 3,991 | 78.1 |  |
| POS | 123 | 2.4 | 121 | 2.4 |  |
| Indemnity | 233 | 4.6 | 228 | 4.5 |  |
| Other/Unknown | 64 | 1.3 | 69 | 1.4 |  |
| Charlson comorbidity index categories |  |  |  |  |  |
| 0 | 609 | 11.9 | 599 | 11.7 | 0.02 |
| 1 | 765 | 15.0 | 794 | 15.5 |  |
| 2 | 866 | 16.9 | 875 | 17.1 |  |
| 3 | 756 | 14.8 | 764 | 14.9 |  |
| ≥4 | 2,115 | 41.4 | 2,079 | 40.7 |  |
| Specific comorbidities |  |  |  |  |  |
| COPD | 855 | 16.7 | 864 | 16.9 | 0.005 |
| Chronic pulmonary disease | 174 | 3.4 | 169 | 3.3 | -0.01 |
| Asthma | 319 | 6.2 | 284 | 5.6 | -0.03 |
| CHF | 1036 | 20.3 | 1041 | 20.4 | 0.00 |
| Valvular disease | 570 | 11.2 | 596 | 11.7 | 0.02 |
| CAD | 5111 | 100.0 | 5111 | 100.0 | 0.00 |
| Old MI | 344 | 6.7 | 325 | 6.4 | -0.02 |
| Acute MI | 361 | 7.1 | 369 | 7.2 | 0.01 |
| Stroke | 487 | 9.5 | 475 | 9.3 | -0.01 |
| Atherosclerosis | 168 | 3.3 | 172 | 3.4 | 0.004 |
| Early stage CKD | 641 | 12.5 | 663 | 13.0 | 0.01 |
| Late stage CKD | 185 | 3.6 | 186 | 3.6 | 0.001 |
| Influenza vaccination in the index influenza season before the index date | 1,502 | 29.4 | 1,467 | 28.7 | -0.02 |
| Index influenza season |  |  |  |  |  |
| 2014 (Oct 2014 - May 2015) | 1,532 | 30.0 | 1,494 | 29.2 | 0.02 |
| 2015 (Oct 2015 - May 2016) | 471 | 9.2 | 488 | 9.5 |  |
| 2016 (Oct 2016 - May 2017) | 1,131 | 22.1 | 1,147 | 22.4 |  |
| 2017 (Oct 2017 - May 2018) | 1,573 | 30.8 | 1,569 | 30.7 |  |
| 2018 (Oct 2018 - March 2019) | 404 | 7.9 | 413 | 8.1 |  |
| Index month |  |  |  |  |  |
| October | 74 | 1.4 | 70 | 1.4 | 0.03 |
| November | 155 | 3.0 | 155 | 3.0 |  |
| December | 853 | 16.7 | 834 | 16.3 |  |
| January | 1,555 | 30.4 | 1,575 | 30.8 |  |
| February | 1,412 | 27.6 | 1,414 | 27.7 |  |
| March | 700 | 13.7 | 688 | 13.5 |  |
| April | 280 | 5.5 | 275 | 5.4 |  |
| May | 82 | 1.6 | 100 | 2.0 |  |
| Total all-cause healthcare costs during the 12-month baseline period ($) |  |  |  |  |  |
| Mean (SD) | $43,612 | $95,064 | $46,123 | $98,097 | -0.01 |
| * Demographic measures (age, sex, payer type, plan type, and geographic region) and timing of index date (index influenza season and index month) were reported from data on the index date. Baseline clinical characteristics (Charlson comorbidity index, specific comorbidities, evidence of influenza vaccine, and baseline total healthcare costs) were reported using data from the 12-month baseline period. | | | | | |

### Supplementary Table 7. Baseline demographic and clinical characteristics for influenza and non-influenza patients with baseline congestive heart failure (CHF) after PS-matching

|  | CHF | | | | |
| --- | --- | --- | --- | --- | --- |
| Measures* | Influenza cohort | | Non-influenza cohort | | Std. Diff. |
|  | N=1,930 | | N=1,930 | |  |
|  | N | % | N | % |  |
| Age categories, years |  |  |  |  |  |
| 65-74 | 884 | 45.8 | 884 | 45.8 | 0.00 |
| 75+ | 1,046 | 54.2 | 1,046 | 54.2 |  |
| Sex |  |  |  |  |  |
| Male | 1,022 | 53.0 | 1,022 | 53.0 | 0.00 |
| Female | 908 | 47.0 | 908 | 47.0 |  |
| Geographic region |  |  |  |  |  |
| Northeast | 534 | 27.7 | 525 | 27.2 | 0.06 |
| Midwest | 321 | 16.6 | 309 | 16.0 |  |
| South | 571 | 29.6 | 538 | 27.9 |  |
| West | 504 | 26.1 | 558 | 28.9 |  |
| Payer type |  |  |  |  |  |
| Commercial | 768 | 39.8 | 753 | 39.0 | 0.05 |
| Self-insured | 562 | 29.1 | 549 | 28.4 |  |
| Medicaid | 74 | 3.8 | 65 | 3.4 |  |
| Medicare risk | 516 | 26.7 | 550 | 28.5 |  |
| Other/Unknown | 10 | 0.5 | 13 | 0.7 |  |
| Plan type |  |  |  |  |  |
| HMO | 452 | 23.4 | 464 | 24.0 | 0.05 |
| PPO | 1,310 | 67.9 | 1,275 | 66.1 |  |
| POS | 28 | 1.5 | 33 | 1.7 |  |
| Indemnity | 119 | 6.2 | 131 | 6.8 |  |
| Other/Unknown | 21 | 1.1 | 27 | 1.4 |  |
| Charlson comorbidity index categories |  |  |  |  |  |
| 0 | 0 | 0.0 | 0 | 0.0 | 0.01 |
| 1 | 86 | 4.5 | 87 | 4.5 |  |
| 2 | 199 | 10.3 | 191 | 9.9 |  |
| 3 | 252 | 13.1 | 253 | 13.1 |  |
| ≥4 | 1,393 | 72.2 | 1,399 | 72.5 |  |
| Specific comorbidities |  |  |  |  |  |
| COPD | 600 | 31.1 | 586 | 30.4 | -0.02 |
| Chronic pulmonary disease | 110 | 5.7 | 105 | 5.4 | -0.01 |
| Asthma | 203 | 10.5 | 194 | 10.1 | -0.02 |
| CHF | 1,930 | 100.0 | 1,930 | 100.0 | 0.00 |
| Valvular disease | 431 | 22.3 | 439 | 22.7 | 0.01 |
| CAD | 1,036 | 53.7 | 998 | 51.7 | -0.04 |
| Old MI | 122 | 6.3 | 123 | 6.4 | 0.002 |
| Acute MI | 215 | 11.1 | 196 | 10.2 | -0.03 |
| Stroke | 242 | 12.5 | 221 | 11.5 | -0.03 |
| Atherosclerosis | 99 | 5.1 | 95 | 4.9 | -0.01 |
| Early stage CKD | 500 | 25.9 | 514 | 26.6 | 0.02 |
| Late stage CKD | 174 | 9.0 | 183 | 9.5 | 0.02 |
| Influenza vaccination in the index influenza season before the index date | 585 | 30.3 | 570 | 29.5 | -0.02 |
| Index influenza season |  |  |  |  |  |
| 2014 (Oct 2014 - May 2015) | 563 | 29.2 | 535 | 27.7 | 0.05 |
| 2015 (Oct 2015 - May 2016) | 188 | 9.7 | 198 | 10.3 |  |
| 2016 (Oct 2016 - May 2017) | 456 | 23.6 | 477 | 24.7 |  |
| 2017 (Oct 2017 - May 2018) | 574 | 29.7 | 583 | 30.2 |  |
| 2018 (Oct 2018 - March 2019) | 149 | 7.7 | 137 | 7.1 |  |
| Index month |  |  |  |  |  |
| October | 25 | 1.3 | 21 | 1.1 | 0.04 |
| November | 59 | 3.1 | 58 | 3.0 |  |
| December | 343 | 17.8 | 359 | 18.6 |  |
| January | 594 | 30.8 | 588 | 30.5 |  |
| February | 504 | 26.1 | 490 | 25.4 |  |
| March | 258 | 13.4 | 276 | 14.3 |  |
| April | 113 | 5.9 | 108 | 5.6 |  |
| May | 34 | 1.8 | 30 | 1.6 |  |
| Total all-cause healthcare costs during the 12-month baseline period ($) |  |  |  |  |  |
| Mean (SD) | $76,425 | $152,502 | $70,982 | $124,187 | -0.01 |
| * Demographic measures (age, sex, payer type, plan type, and geographic region) and timing of index date (index influenza season and index month) were reported from data on the index date. Baseline clinical characteristics (Charlson comorbidity index, specific comorbidities, evidence of influenza vaccine, and baseline total healthcare costs) were reported using data from the 12-month baseline period. | | | | | |

### Supplementary Table 8. Baseline demographic and clinical characteristics for influenza and non-influenza patients with baseline stroke after PS-matching

|  | Stroke | | | | |
| --- | --- | --- | --- | --- | --- |
| Measures* | Influenza cohort | | Non-influenza cohort | | Std. Diff. |
|  | N=1,287 | | N=1,287 | |  |
|  | N | % | N | % |  |
| Age categories, years |  |  |  |  |  |
| 65-74 | 724 | 56.3 | 724 | 56.3 | 0.00 |
| 75+ | 563 | 43.7 | 563 | 43.7 |  |
| Sex |  |  |  |  |  |
| Male | 678 | 52.7 | 678 | 52.7 | 0.00 |
| Female | 609 | 47.3 | 609 | 47.3 |  |
| Geographic region |  |  |  |  |  |
| Northeast | 303 | 23.5 | 341 | 26.5 | 0.07 |
| Midwest | 202 | 15.7 | 187 | 14.5 |  |
| South | 491 | 38.2 | 470 | 36.5 |  |
| West | 291 | 22.6 | 289 | 22.5 |  |
| Payer type |  |  |  |  |  |
| Commercial | 543 | 42.2 | 547 | 42.5 | 0.05 |
| Self-insured | 464 | 36.1 | 453 | 35.2 |  |
| Medicaid | 30 | 2.3 | 23 | 1.8 |  |
| Medicare risk | 242 | 18.8 | 253 | 19.7 |  |
| Other/Unknown | 8 | 0.6 | 11 | 0.9 |  |
| Plan type |  |  |  |  |  |
| HMO | 236 | 18.3 | 221 | 17.2 | 0.06 |
| PPO | 940 | 73.0 | 955 | 74.2 |  |
| POS | 24 | 1.9 | 24 | 1.9 |  |
| Indemnity | 74 | 5.7 | 68 | 5.3 |  |
| Other/Unknown | 13 | 1.0 | 19 | 1.5 |  |
| Charlson comorbidity index categories |  |  |  |  |  |
| 0 | 13 | 1.0 | 15 | 1.2 | 0.03 |
| 1 | 152 | 11.8 | 163 | 12.7 |  |
| 2 | 193 | 15.0 | 187 | 14.5 |  |
| 3 | 193 | 15.0 | 191 | 14.8 |  |
| ≥4 | 736 | 57.2 | 731 | 56.8 |  |
| Specific comorbidities |  |  |  |  |  |
| COPD | 239 | 18.6 | 249 | 19.3 | 0.02 |
| Chronic pulmonary disease | 40 | 3.1 | 48 | 3.7 | 0.03 |
| Asthma | 102 | 7.9 | 103 | 8.0 | 0.003 |
| CHF | 242 | 18.8 | 237 | 18.4 | -0.01 |
| Valvular disease | 155 | 12.0 | 156 | 12.1 | 0.00 |
| CAD | 487 | 37.8 | 497 | 38.6 | 0.02 |
| Old MI | 61 | 4.7 | 55 | 4.3 | -0.02 |
| Acute MI | 74 | 5.7 | 67 | 5.2 | -0.02 |
| Stroke | 1,287 | 100.0 | 1,287 | 100.0 | 0.00 |
| Atherosclerosis | 64 | 5.0 | 72 | 5.6 | 0.03 |
| Early stage CKD | 181 | 14.1 | 165 | 12.8 | -0.04 |
| Late stage CKD | 47 | 3.7 | 37 | 2.9 | -0.04 |
| Influenza vaccination in the index influenza season before the index date | 352 | 27.4 | 349 | 27.1 | -0.01 |
| Index influenza season |  |  |  |  |  |
| 2014 (Oct 2014 - May 2015) | 416 | 32.3 | 426 | 33.1 | 0.04 |
| 2015 (Oct 2015 - May 2016) | 132 | 10.3 | 121 | 9.4 |  |
| 2016 (Oct 2016 - May 2017) | 289 | 22.5 | 295 | 22.9 |  |
| 2017 (Oct 2017 - May 2018) | 361 | 28.0 | 352 | 27.4 |  |
| 2018 (Oct 2018 - March 2019) | 89 | 6.9 | 93 | 7.2 |  |
| Index month |  |  |  |  |  |
| October | 26 | 2.0 | 18 | 1.4 | 0.10 |
| November | 34 | 2.6 | 45 | 3.5 |  |
| December | 240 | 18.6 | 265 | 20.6 |  |
| January | 387 | 30.1 | 372 | 28.9 |  |
| February | 339 | 26.3 | 343 | 26.7 |  |
| March | 176 | 13.7 | 170 | 13.2 |  |
| April | 72 | 5.6 | 57 | 4.4 |  |
| May | 13 | 1.0 | 17 | 1.3 |  |
| Total all-cause healthcare costs during the 12-month baseline period ($) |  |  |  |  |  |
| Mean (SD) | $63,760 | $162,100 | $55,376 | $84,295 | -0.004 |
| * Demographic measures (age, sex, payer type, plan type, and geographic region) and timing of index date (index influenza season and index month) were reported from data on the index date. Baseline clinical characteristics (Charlson comorbidity index, specific comorbidities, evidence of influenza vaccine, and baseline total healthcare costs) were reported using data from the 12-month baseline period. | | | | | |

### Supplementary Table 9. Baseline demographic and clinical characteristics for influenza and non-influenza patients with baseline valvular disease after PS-matching

|  | Valvular disease | | | | |
| --- | --- | --- | --- | --- | --- |
| Measures* | Influenza cohort | | Non-influenza cohort | | Std. Diff. |
|  | N=1,380 | | N=1,380 | |  |
|  | N | % | N | % |  |
| Age categories, years |  |  |  |  |  |
| 65-74 | 736 | 53.3 | 736 | 53.3 | 0.00 |
| 75+ | 644 | 46.7 | 644 | 46.7 |  |
| Sex |  |  |  |  |  |
| Male | 708 | 51.3 | 708 | 51.3 | 0.00 |
| Female | 672 | 48.7 | 672 | 48.7 |  |
| Geographic region |  |  |  |  |  |
| Northeast | 377 | 27.3 | 390 | 28.3 | 0.06 |
| Midwest | 199 | 14.4 | 183 | 13.3 |  |
| South | 501 | 36.3 | 528 | 38.3 |  |
| West | 303 | 22.0 | 279 | 20.2 |  |
| Payer type |  |  |  |  |  |
| Commercial | 624 | 45.2 | 629 | 45.6 | 0.05 |
| Self-insured | 458 | 33.2 | 456 | 33.0 |  |
| Medicaid | 28 | 2.0 | 22 | 1.6 |  |
| Medicare risk | 256 | 18.6 | 254 | 18.4 |  |
| Other/Unknown | 14 | 1.0 | 19 | 1.4 |  |
| Plan type |  |  |  |  |  |
| HMO | 213 | 15.4 | 215 | 15.6 | 0.04 |
| PPO | 1,047 | 75.9 | 1,049 | 76.0 |  |
| POS | 20 | 1.4 | 15 | 1.1 |  |
| Indemnity | 81 | 5.9 | 79 | 5.7 |  |
| Other/Unknown | 19 | 1.4 | 22 | 1.6 |  |
| Charlson comorbidity index categories |  |  |  |  |  |
| 0 | 164 | 11.9 | 169 | 12.2 | 0.07 |
| 1 | 202 | 14.6 | 171 | 12.4 |  |
| 2 | 224 | 16.2 | 242 | 17.5 |  |
| 3 | 184 | 13.3 | 182 | 13.2 |  |
| ≥4 | 606 | 43.9 | 616 | 44.6 |  |
| Specific comorbidities |  |  |  |  |  |
| COPD | 234 | 17.0 | 238 | 17.2 | 0.01 |
| Chronic pulmonary disease | 48 | 3.5 | 45 | 3.3 | -0.01 |
| Asthma | 132 | 9.6 | 139 | 10.1 | 0.02 |
| CHF | 431 | 31.2 | 432 | 31.3 | 0.00 |
| Valvular disease | 1380 | 100.0 | 1380 | 100.0 | 0.00 |
| CAD | 570 | 41.3 | 592 | 42.9 | 0.03 |
| Old MI | 55 | 4.0 | 63 | 4.6 | 0.03 |
| Acute MI | 77 | 5.6 | 88 | 6.4 | 0.03 |
| Stroke | 155 | 11.2 | 157 | 11.4 | 0.005 |
| Atherosclerosis | 58 | 4.2 | 66 | 4.8 | 0.03 |
| Early stage CKD | 212 | 15.4 | 206 | 14.9 | -0.01 |
| Late stage CKD | 50 | 3.6 | 45 | 3.3 | -0.02 |
| Influenza vaccination in the index influenza season before the index date | 448 | 32.5 | 460 | 33.3 | 0.02 |
| Index influenza season |  |  |  |  |  |
| 2014 (Oct 2014 - May 2015) | 376 | 27.2 | 382 | 27.7 | 0.04 |
| 2015 (Oct 2015 - May 2016) | 131 | 9.5 | 125 | 9.1 |  |
| 2016 (Oct 2016 - May 2017) | 342 | 24.8 | 354 | 25.7 |  |
| 2017 (Oct 2017 - May 2018) | 418 | 30.3 | 417 | 30.2 |  |
| 2018 (Oct 2018 - March 2019) | 113 | 8.2 | 102 | 7.4 |  |
| Index month |  |  |  |  |  |
| October | 14 | 1.0 | 13 | 0.9 | 0.06 |
| November | 41 | 3.0 | 45 | 3.3 |  |
| December | 212 | 15.4 | 215 | 15.6 |  |
| January | 435 | 31.5 | 419 | 30.4 |  |
| February | 365 | 26.4 | 357 | 25.9 |  |
| March | 194 | 14.1 | 218 | 15.8 |  |
| April | 95 | 6.9 | 89 | 6.4 |  |
| May | 24 | 1.7 | 24 | 1.7 |  |
| Total all-cause healthcare costs during the 12-month baseline period ($) |  |  |  |  |  |
| Mean (SD) | $56,941 | $152,927 | $53,311 | $106,937 | 0.002 |
| * Demographic measures (age, sex, payer type, plan type, and geographic region) and timing of index date (index influenza season and index month) were reported from data on the index date. Baseline clinical characteristics (Charlson comorbidity index, specific comorbidities, evidence of influenza vaccine, and baseline total healthcare costs) were reported using data from the 12-month baseline period. | | | | | |

### Supplementary Table 10. Baseline demographic and clinical characteristics for influenza and non-influenza patients with history of old myocardial infarction (MI) after PS-matching

|  | Old MI | | | | |
| --- | --- | --- | --- | --- | --- |
| Measures* | Influenza cohort | | Non-influenza cohort | | Std. Diff. |
|  | N=397 | | N=397 | |  |
|  | N | % | N | % |  |
| Age categories, years |  |  |  |  |  |
| 65-74 | 261 | 65.7 | 261 | 65.7 | 0.00 |
| 75+ | 136 | 34.3 | 136 | 34.3 |  |
| Sex |  |  |  |  |  |
| Male | 271 | 68.3 | 271 | 68.3 | 0.00 |
| Female | 126 | 31.7 | 126 | 31.7 |  |
| Geographic region |  |  |  |  |  |
| Northeast | 75 | 18.9 | 71 | 17.9 | 0.09 |
| Midwest | 78 | 19.6 | 69 | 17.4 |  |
| South | 136 | 34.3 | 151 | 38.0 |  |
| West | 108 | 27.2 | 106 | 26.7 |  |
| Payer type |  |  |  |  |  |
| Commercial | 161 | 40.6 | 169 | 42.6 | 0.08 |
| Self-insured | 152 | 38.3 | 152 | 38.3 |  |
| Medicaid | 15 | 3.8 | 11 | 2.8 |  |
| Medicare risk | 67 | 16.9 | 64 | 16.1 |  |
| Other/Unknown | 2 | 0.5 | 1 | 0.3 |  |
| Plan type |  |  |  |  |  |
| HMO | 91 | 22.9 | 90 | 22.7 | 0.12 |
| PPO | 270 | 68.0 | 283 | 71.3 |  |
| POS | 7 | 1.8 | 4 | 1.0 |  |
| Indemnity | 28 | 7.1 | 19 | 4.8 |  |
| Other/Unknown | 1 | 0.3 | 1 | 0.3 |  |
| Charlson comorbidity index categories |  |  |  |  |  |
| 0 | 0 | 0.0 | 0 | 0.0 | 0.09 |
| 1 | 39 | 9.8 | 39 | 9.8 |  |
| 2 | 47 | 11.8 | 44 | 11.1 |  |
| 3 | 66 | 16.6 | 79 | 19.9 |  |
| ≥4 | 245 | 61.7 | 235 | 59.2 |  |
| Specific comorbidities |  |  |  |  |  |
| COPD | 101 | 25.4 | 104 | 26.2 | 0.02 |
| Chronic pulmonary disease | 16 | 4.0 | 13 | 3.3 | -0.04 |
| Asthma | 27 | 6.8 | 26 | 6.5 | -0.01 |
| CHF | 121 | 30.5 | 112 | 28.2 | -0.05 |
| Valvular disease | 54 | 13.6 | 55 | 13.9 | 0.01 |
| CAD | 343 | 86.4 | 348 | 87.7 | 0.04 |
| Old MI | 397 | 100.0 | 397 | 100.0 | 0.00 |
| Acute MI | 0 | 0.0 | 0 | 0.0 | 0.00 |
| Stroke | 61 | 15.4 | 54 | 13.6 | -0.05 |
| Atherosclerosis | 19 | 4.8 | 24 | 6.0 | 0.06 |
| Early stage CKD | 74 | 18.6 | 73 | 18.4 | -0.01 |
| Late stage CKD | 25 | 6.3 | 33 | 8.3 | 0.08 |
| Influenza vaccination in the index influenza season before the index date | 125 | 31.5 | 131 | 33.0 | 0.03 |
| Index influenza season |  |  |  |  |  |
| 2014 (Oct 2014 - May 2015) | 111 | 28.0 | 113 | 28.5 | 0.06 |
| 2015 (Oct 2015 - May 2016) | 36 | 9.1 | 36 | 9.1 |  |
| 2016 (Oct 2016 - May 2017) | 91 | 22.9 | 89 | 22.4 |  |
| 2017 (Oct 2017 - May 2018) | 119 | 30.0 | 113 | 28.5 |  |
| 2018 (Oct 2018 - March 2019) | 40 | 10.1 | 46 | 11.6 |  |
| Index month |  |  |  |  |  |
| October | 8 | 2.0 | 8 | 2.0 | 0.13 |
| November | 6 | 1.5 | 9 | 2.3 |  |
| December | 53 | 13.4 | 64 | 16.1 |  |
| January | 115 | 29.0 | 104 | 26.2 |  |
| February | 133 | 33.5 | 122 | 30.7 |  |
| March | 50 | 12.6 | 52 | 13.1 |  |
| April | 22 | 5.5 | 28 | 7.1 |  |
| May | 10 | 2.5 | 10 | 2.5 |  |
| Total all-cause healthcare costs during the 12-month baseline period ($) |  |  |  |  |  |
| Mean (SD) | $64,294 | $136,991 | $69,161 | $114,137 | 0.04 |
| * Demographic measures (age, sex, payer type, plan type, and geographic region) and timing of index date (index influenza season and index month) were reported from data on the index date. Baseline clinical characteristics (Charlson comorbidity index, specific comorbidities, evidence of influenza vaccine, and baseline total healthcare costs) were reported using data from the 12-month baseline period. | | | | | |

### Supplementary Table 11. Baseline demographic and clinical characteristics for influenza and non-influenza patients with baseline acute MI after PS-matching

|  | Acute MI | | | | |
| --- | --- | --- | --- | --- | --- |
| Measures* | Influenza cohort | | Non-influenza cohort | | Std. Diff. |
|  | N=473 | | N=473 | |  |
|  | N | % | N | % |  |
| Age categories, years |  |  |  |  |  |
| 65-74 | 272 | 57.5 | 272 | 57.5 | 0.00 |
| 75+ | 201 | 42.5 | 201 | 42.5 |  |
| Sex |  |  |  |  |  |
| Male | 298 | 63.0 | 298 | 63.0 | 0.00 |
| Female | 175 | 37.0 | 175 | 37.0 |  |
| Geographic region |  |  |  |  |  |
| Northeast | 155 | 32.8 | 154 | 32.6 | 0.08 |
| Midwest | 76 | 16.1 | 82 | 17.3 |  |
| South | 127 | 26.8 | 113 | 23.9 |  |
| West | 115 | 24.3 | 124 | 26.2 |  |
| Payer type |  |  |  |  |  |
| Commercial | 216 | 45.7 | 204 | 43.1 | 0.11 |
| Self-insured | 136 | 28.8 | 139 | 29.4 |  |
| Medicaid | 8 | 1.7 | 10 | 2.1 |  |
| Medicare risk | 111 | 23.5 | 120 | 25.4 |  |
| Other/Unknown | 2 | 0.4 | 0 | 0.0 |  |
| Plan type |  |  |  |  |  |
| HMO | 100 | 21.1 | 106 | 22.4 | 0.13 |
| PPO | 332 | 70.2 | 325 | 68.7 |  |
| POS | 14 | 3.0 | 12 | 2.5 |  |
| Indemnity | 24 | 5.1 | 30 | 6.3 |  |
| Other/Unknown | 3 | 0.6 | 0 | 0.0 |  |
| Charlson comorbidity index categories |  |  |  |  |  |
| 0 | 0 | 0.0 | 0 | 0.0 | 0.03 |
| 1 | 39 | 8.2 | 35 | 7.4 |  |
| 2 | 56 | 11.8 | 58 | 12.3 |  |
| 3 | 52 | 11.0 | 51 | 10.8 |  |
| ≥4 | 326 | 68.9 | 329 | 69.6 |  |
| Specific comorbidities |  |  |  |  |  |
| COPD | 119 | 25.2 | 121 | 25.6 | 0.01 |
| Chronic pulmonary disease | 24 | 5.1 | 13 | 2.7 | -0.12 |
| Asthma | 46 | 9.7 | 48 | 10.1 | 0.01 |
| CHF | 211 | 44.6 | 223 | 47.1 | 0.05 |
| Valvular disease | 75 | 15.9 | 80 | 16.9 | 0.03 |
| CAD | 357 | 75.5 | 347 | 73.4 | -0.05 |
| Old MI | 0 | 0.0 | 0 | 0.0 | 0.00 |
| Acute MI | 473 | 100.0 | 473 | 100.0 | 0.00 |
| Stroke | 72 | 15.2 | 68 | 14.4 | -0.02 |
| Atherosclerosis | 28 | 5.9 | 30 | 6.3 | 0.02 |
| Early stage CKD | 95 | 20.1 | 102 | 21.6 | 0.04 |
| Late stage CKD | 43 | 9.1 | 45 | 9.5 | 0.01 |
| Influenza vaccination in the index influenza season before the index date | 140 | 29.6 | 144 | 30.4 | 0.02 |
| Index influenza season |  |  |  |  |  |
| 2014 (Oct 2014 - May 2015) | 117 | 24.7 | 124 | 26.2 | 0.04 |
| 2015 (Oct 2015 - May 2016) | 54 | 11.4 | 51 | 10.8 |  |
| 2016 (Oct 2016 - May 2017) | 113 | 23.9 | 112 | 23.7 |  |
| 2017 (Oct 2017 - May 2018) | 152 | 32.1 | 148 | 31.3 |  |
| 2018 (Oct 2018 - March 2019) | 37 | 7.8 | 38 | 8.0 |  |
| Index month |  |  |  |  |  |
| October | 7 | 1.5 | 9 | 1.9 | 0.07 |
| November | 12 | 2.5 | 13 | 2.7 |  |
| December | 76 | 16.1 | 79 | 16.7 |  |
| January | 144 | 30.4 | 139 | 29.4 |  |
| February | 119 | 25.2 | 111 | 23.5 |  |
| March | 74 | 15.6 | 75 | 15.9 |  |
| April | 34 | 7.2 | 38 | 8.0 |  |
| May | 7 | 1.5 | 9 | 1.9 |  |
| Total all-cause healthcare costs during the 12-month baseline period ($) |  |  |  |  |  |
| Mean (SD) | $96,136 | $184,831 | $95,129 | $131,197 | 0.04 |
| * Demographic measures (age, sex, payer type, plan type, and geographic region) and timing of index date (index influenza season and index month) were reported from data on the index date. Baseline clinical characteristics (Charlson comorbidity index, specific comorbidities, evidence of influenza vaccine, and baseline total healthcare costs) were reported using data from the 12-month baseline period. | | | | | |

### Supplementary Table 12. Baseline demographic and clinical characteristics for influenza and non-influenza patients with baseline early stage chronic kidney disease (CKD) after PS-matching

|  | Early stage CKD | | | | |
| --- | --- | --- | --- | --- | --- |
| Measures* | Influenza cohort | | Non-influenza cohort | | Std. Diff. |
|  | N=2,020 | | N=2,020 | |  |
|  | N | % | N | % |  |
| Age categories, years |  |  |  |  |  |
| 65-74 | 1,062 | 52.6 | 1,062 | 52.6 | 0.00 |
| 75+ | 958 | 47.4 | 958 | 47.4 |  |
| Sex |  |  |  |  |  |
| Male | 1,106 | 54.8 | 1,106 | 54.8 | 0.00 |
| Female | 914 | 45.2 | 914 | 45.2 |  |
| Geographic region |  |  |  |  |  |
| Northeast | 430 | 21.3 | 437 | 21.6 | 0.03 |
| Midwest | 348 | 17.2 | 355 | 17.6 |  |
| South | 690 | 34.2 | 700 | 34.7 |  |
| West | 552 | 27.3 | 528 | 26.1 |  |
| Payer type |  |  |  |  |  |
| Commercial | 916 | 45.3 | 904 | 44.8 | 0.02 |
| Self-insured | 676 | 33.5 | 684 | 33.9 |  |
| Medicaid | 63 | 3.1 | 59 | 2.9 |  |
| Medicare risk | 353 | 17.5 | 360 | 17.8 |  |
| Other/Unknown | 12 | 0.6 | 13 | 0.6 |  |
| Plan type |  |  |  |  |  |
| HMO | 386 | 19.1 | 379 | 18.8 | 0.04 |
| PPO | 1,447 | 71.6 | 1,434 | 71.0 |  |
| POS | 38 | 1.9 | 39 | 1.9 |  |
| Indemnity | 126 | 6.2 | 139 | 6.9 |  |
| Other/Unknown | 23 | 1.1 | 29 | 1.4 |  |
| Charlson comorbidity index categories |  |  |  |  |  |
| 0 | 5 | 0.2 | 3 | 0.1 | 0.06 |
| 1 | 1 | 0.0 | 3 | 0.1 |  |
| 2 | 197 | 9.8 | 197 | 9.8 |  |
| 3 | 257 | 12.7 | 228 | 11.3 |  |
| ≥4 | 1,560 | 77.2 | 1,589 | 78.7 |  |
| Specific comorbidities |  |  |  |  |  |
| COPD | 327 | 16.2 | 332 | 16.4 | 0.01 |
| Chronic pulmonary disease | 79 | 3.9 | 68 | 3.4 | -0.03 |
| Asthma | 176 | 8.7 | 162 | 8.0 | -0.03 |
| CHF | 499 | 24.7 | 524 | 25.9 | 0.03 |
| Valvular disease | 212 | 10.5 | 211 | 10.4 | -0.002 |
| CAD | 640 | 31.7 | 679 | 33.6 | 0.04 |
| Old MI | 75 | 3.7 | 81 | 4.0 | 0.02 |
| Acute MI | 94 | 4.7 | 95 | 4.7 | 0.002 |
| Stroke | 181 | 9.0 | 203 | 10.0 | 0.04 |
| Atherosclerosis | 84 | 4.2 | 70 | 3.5 | -0.04 |
| Early stage CKD | 2,020 | 100.0 | 2,020 | 100.0 | 0.00 |
| Late stage CKD | 0 | 0.0 | 0 | 0.0 | 0.00 |
| Influenza vaccination in the index influenza season before the index date | 609 | 30.1 | 614 | 30.4 | 0.01 |
| Index influenza season |  |  |  |  |  |
| 2014 (Oct 2014 - May 2015) | 539 | 26.7 | 537 | 26.6 | 0.03 |
| 2015 (Oct 2015 - May 2016) | 204 | 10.1 | 217 | 10.7 |  |
| 2016 (Oct 2016 - May 2017) | 499 | 24.7 | 482 | 23.9 |  |
| 2017 (Oct 2017 - May 2018) | 619 | 30.6 | 630 | 31.2 |  |
| 2018 (Oct 2018 - March 2019) | 159 | 7.9 | 154 | 7.6 |  |
| Index month |  |  |  |  |  |
| October | 31 | 1.5 | 39 | 1.9 | 0.04 |
| November | 57 | 2.8 | 51 | 2.5 |  |
| December | 336 | 16.6 | 338 | 16.7 |  |
| January | 599 | 29.7 | 600 | 29.7 |  |
| February | 516 | 25.5 | 513 | 25.4 |  |
| March | 291 | 14.4 | 283 | 14.0 |  |
| April | 146 | 7.2 | 145 | 7.2 |  |
| May | 44 | 2.2 | 51 | 2.5 |  |
| Total all-cause healthcare costs during the 12-month baseline period ($) |  |  |  |  |  |
| Mean (SD) | $49,206 | $96,554 | $52,249 | $118,407 | 0.02 |
| * Demographic measures (age, sex, payer type, plan type, and geographic region) and timing of index date (index influenza season and index month) were reported from data on the index date. Baseline clinical characteristics (Charlson comorbidity index, specific comorbidities, evidence of influenza vaccine, and baseline total healthcare costs) were reported using data from the 12-month baseline period. | | | | | |

### Supplementary Table 13. Baseline demographic and clinical characteristics for influenza and non-influenza patients with baseline late stage CKD after PS-matching

|  | Late stage CKD | | | | |
| --- | --- | --- | --- | --- | --- |
| Measures* | Influenza cohort | | Non-influenza cohort | | Std. Diff. |
|  | N=429 | | N=429 | |  |
|  | N | % | N | % |  |
| Age categories, years |  |  |  |  |  |
| 65-74 | 271 | 63.2 | 271 | 63.2 | 0.00 |
| 75+ | 158 | 36.8 | 158 | 36.8 |  |
| Sex |  |  |  |  |  |
| Male | 252 | 58.7 | 252 | 58.7 | 0.00 |
| Female | 177 | 41.3 | 177 | 41.3 |  |
| Geographic region |  |  |  |  |  |
| Northeast | 107 | 24.9 | 102 | 23.8 | 0.04 |
| Midwest | 94 | 21.9 | 91 | 21.2 |  |
| South | 126 | 29.4 | 132 | 30.8 |  |
| West | 102 | 23.8 | 104 | 24.2 |  |
| Payer type |  |  |  |  |  |
| Commercial | 206 | 48.0 | 207 | 48.3 | 0.06 |
| Self-insured | 132 | 30.8 | 132 | 30.8 |  |
| Medicaid | 18 | 4.2 | 20 | 4.7 |  |
| Medicare risk | 69 | 16.1 | 64 | 14.9 |  |
| Other/Unknown | 4 | 0.9 | 6 | 1.4 |  |
| Plan type |  |  |  |  |  |
| HMO | 71 | 16.6 | 73 | 17.0 | 0.04 |
| PPO | 320 | 74.6 | 314 | 73.2 |  |
| POS | 12 | 2.8 | 13 | 3.0 |  |
| Indemnity | 21 | 4.9 | 23 | 5.4 |  |
| Other/Unknown | 5 | 1.2 | 6 | 1.4 |  |
| Charlson comorbidity index categories |  |  |  |  |  |
| 0 | 0 | 0.0 | 1 | 0.2 | 0.07 |
| 1 | 0 | 0.0 | 0 | 0.0 |  |
| 2 | 23 | 5.4 | 22 | 5.1 |  |
| 3 | 31 | 7.2 | 28 | 6.5 |  |
| ≥4 | 375 | 87.4 | 378 | 88.1 |  |
| Specific comorbidities |  |  |  |  |  |
| COPD | 96 | 22.4 | 87 | 20.3 | -0.05 |
| Chronic pulmonary disease | 23 | 5.4 | 27 | 6.3 | 0.04 |
| Asthma | 26 | 6.1 | 31 | 7.2 | 0.05 |
| CHF | 172 | 40.1 | 171 | 39.9 | -0.005 |
| Valvular disease | 49 | 11.4 | 44 | 10.3 | -0.04 |
| CAD | 183 | 42.7 | 170 | 39.6 | -0.06 |
| Old MI | 24 | 5.6 | 21 | 4.9 | -0.03 |
| Acute MI | 45 | 10.5 | 53 | 12.4 | 0.06 |
| Stroke | 46 | 10.7 | 43 | 10.0 | -0.02 |
| Atherosclerosis | 28 | 6.5 | 29 | 6.8 | 0.01 |
| Early stage CKD | 0 | 0.0 | 0 | 0.0 | 0.00 |
| Late stage CKD | 429 | 100.0 | 429 | 100.0 | 0.00 |
| Influenza vaccination in the index influenza season before the index date | 147 | 34.3 | 152 | 35.4 | 0.02 |
| Index influenza season |  |  |  |  |  |
| 2014 (Oct 2014 - May 2015) | 133 | 31.0 | 142 | 33.1 | 0.11 |
| 2015 (Oct 2015 - May 2016) | 50 | 11.7 | 49 | 11.4 |  |
| 2016 (Oct 2016 - May 2017) | 93 | 21.7 | 94 | 21.9 |  |
| 2017 (Oct 2017 - May 2018) | 126 | 29.4 | 109 | 25.4 |  |
| 2018 (Oct 2018 - March 2019) | 27 | 6.3 | 35 | 8.2 |  |
| Index month |  |  |  |  |  |
| October | 5 | 1.2 | 5 | 1.2 | 0.13 |
| November | 10 | 2.3 | 5 | 1.2 |  |
| December | 61 | 14.2 | 71 | 16.6 |  |
| January | 155 | 36.1 | 162 | 37.8 |  |
| February | 118 | 27.5 | 117 | 27.3 |  |
| March | 49 | 11.4 | 41 | 9.6 |  |
| April | 22 | 5.1 | 19 | 4.4 |  |
| May | 9 | 2.1 | 9 | 2.1 |  |
| Total all-cause healthcare costs during the 12-month baseline period ($) |  |  |  |  |  |
| Mean (SD) | $157,833 | $254,925 | $196,106 | $296,100 | 0.04 |
| * Demographic measures (age, sex, payer type, plan type, and geographic region) and timing of index date (index influenza season and index month) were reported from data on the index date. Baseline clinical characteristics (Charlson comorbidity index, specific comorbidities, evidence of influenza vaccine, and baseline total healthcare costs) were reported using data from the 12-month baseline period. | | | | | |
